# RAG-mediated structural variation and its impact on relapse risk in acute lymphoblastic leukemia

**DOI:** 10.64898/2026.05.21.26353542

**Authors:** Tanxin Liu, Yunqi Li, Charley Wang, Cassandra J. Clark, Nathan Anderson, Erin Marcotte, Michael R. Lieber, Srividya Swaminathan, Joseph L. Wiemels, Logan G. Spector, Vijay G. Sankaran, Carol Fries, Adam J. de Smith

**Affiliations:** Center for Genetic Epidemiology, Department of Population and Public Health Sciences, Keck School of Medicine of the University of Southern California, Los Angeles, CA, 90033, USA; Division of Epidemiology and Clinical Research, Department of Pediatrics, University of Minnesota School of Medicine, Minneapolis, MN, 55454, USA; Departments of Pathology, Biochemistry, Molecular Microbiology & Immunology, and Section of Molecular & Computational Biology (Department of Biological Sciences), University of Southern California, Los Angeles, CA, 90033, USA; Department of Systems Biology, Beckman Research Institute of the City of Hope, Monrovia, CA, 91016, USA; Division of Hematology/Oncology, Boston Children’s Hospital and Department of Pediatric Oncology, Dana-Farber Cancer Institute, Harvard Medical School, Boston, MA 02115, USA; Howard Hughes Medical Institute, Boston, MA 02115; Department of Pediatrics, Hematology and Oncology, University of Rochester, Rochester, NY, USA; Department of Pediatrics, Beckman Research Institute of the City of Hope, Duarte, CA, 91010, USA

## Abstract

Relapse during treatment of B-cell acute lymphoblastic leukemia (B-ALL) is a harbinger of poor outcomes. Identifying biomarkers for subsequent relapse risk which are detectable at B-ALL diagnosis remains a priority. Off-target recombination-activating gene (RAG)-mediated structural variants (SVs) generate genomic instability that drives leukemogenesis and may underlie treatment resistance. Leveraging sequencing data in 1,496 pediatric B-ALL patients enriched for relapse status (relapse n=532; non-relapse n=964), we characterized RAG-mediated SVs across B-ALL molecular subtypes and examined their association with patient characteristics and their impact on clinical outcomes. Off-target RAG-mediated SVs were overall frequent, particularly in *ETV6::RUNX1*, *ETV6::RUNX1*-like, and Ph-like B-ALL subtypes, while increasing age-at-diagnosis was positively associated with burden of off-target RAG-mediated SVs (P<.001). Off-target RAG-mediated SVs with a recombination signal sequence (RSS) at one breakpoint, a hallmark of off-target RAG activity, were significantly more frequent at diagnosis in patients who subsequently relapsed (P=.001). This association remained significant in multivariable regression analysis (per SV odds ratio [OR]:1.08, 95%CI:1.04-1.12), in minimal residual disease (MRD)-negative patients (OR:1.09, 95%CI:1.04-1.14) and across subtypes. Excluding deletions, MRD-negative *ETV6::RUNX1* patients with ≥3 off-target RAG-mediated SVs had a >3-fold risk of relapse (hazard ratio:3.47, 95% CI:1.86-6.49). RAG-mediated SVs were also associated with relapse risk in T-cell ALL patients. Off-target RAG-mediated SV burden at diagnosis is a risk factor of relapse in pediatric ALL across molecular subtypes and independent of MRD status.

## Introduction

Most children with pediatric B-cell acute lymphoblastic leukemia (B-ALL) can now be cured with contemporary, risk-adapted, CD19-directed chemoimmunotherapy ^1^, but the intensity and toxicity of curative regimens highlight the need for new predictive biomarkers to enable more precise risk stratification. Despite prognostic advances across B-ALL subgroups with the incorporation of blinatumomab ^1–3^, relapsed B-ALL remains a leading cause of cancer-related mortality in children^4, 5^. As future trials focus on minimizing avoidable treatment-related toxicity while preserving favorable outcomes, identifying the biological determinants that govern leukemogenesis and relapse susceptibility has become increasingly critical.

Epidemiological and experimental evidence support a role for early-life infectious exposure in shaping immune development and influencing ALL risk ^6–8^, while genome-wide association studies (GWAS) have identified germline variants that predispose to ALL, particularly implicating genes involved in lymphoid development ^9, 10^. These observations are consistent with the two-hit model of childhood ALL, in which an initiating germline or prenatal lesion is followed by postnatal acquisition of secondary somatic alterations in key driver genes ^6,11^. A major driver of these somatic structural variations in childhood ALL is illegitimate recombination activating gene (RAG)-mediated recombination ^12, 13^. The RAG1 and RAG2 proteins typically function as part of the adaptive immune system to help generate antibody diversity by recombining immunoglobulin heavy chain (IgH) variable (V), diversity (D), and joining (J) genes followed by light chain V-J genes during the maturation of lymphocytes ^14, 15^. RAG proteins recognize recombination signal sequences (RSS) adjacent to VDJ segments at the immunoglobulin (Ig) and T-cell receptor (TCR) gene regions (“on-target”) and induce DNA double-strand breaks followed by joining of gene segments to form the variable exons of immunoglobulins or TCRs ^16^. In addition, the DNA sequence in between the gene segments is typically excised and the RSS joined to form a signal joint on an excised signal circle (ESC) ^17^. ESCs generated by RAG recombination have been speculated to form a complex with RAG proteins and induce dsDNA breaks at RSS or cryptic RSS across the genome, resulting in the formation of structural variants (SVs) ^16, 18^. Off-target RAG recombination has been implicated in the formation of SVs in non-Ig/TCR regions in B-ALL ^12, 19, 20,16, 18^. Further, the presence of ESCs was recently found to be higher at diagnosis among patients with B-ALL who subsequently relapsed compared with non-relapsed patients ^18^. Here, we uncover a potential etiology of this finding via a comprehensive analysis of RAG recombination-mediated structural variation in the largest clinically annotated cohort of molecularly characterized childhood B-ALL ^21^. Our data shed new light on leukemia pathogenesis and disease behavior.

## Materials and Methods

### Study subjects

We leveraged available data from the Molecular Profiling to Predict Responses to Therapy (MP2PRT) study of B-ALL (dbGaP accession number phs002005.v1.p1), which included 1,496 patients enrolled in Children’s Oncology Group (COG) trials ^21^, to investigate the landscape and predictors of RAG-mediated structural variation in childhood B-ALL at diagnosis, and to examine the association between RAG-mediated SVs and patient outcomes (**Figure S1**). Patients included in MP2PRT predominantly had standard-risk B-ALL (age 1-<10; WBC <50x10^3^/μL at diagnosis) and were enriched for patients who experienced relapse (relapse n=439, non-relapse n=1,057). Raw sequencing data and structural variant (SV) calls for the MP2PRT B-ALL study were available via the National Cancer Institute Genomics Data Commons (GDC) Data Portal through an approved dbGaP request. Patient demographics and other characteristics available in Chang et al.^21^ included age-at-diagnosis, sex, race, ethnicity, molecular subtypes, relapse status, end of induction minimal residual disease (MRD), as well as time-to event data including event-free survival, disease-free survival, and overall survival. Additional details of patient characteristics are described in the Supplemental Methods. This study was reviewed and approved by the Institutional Review Boards at the University of Southern California and City of Hope.

### Sequencing data and SV breakpoint analysis

SV calls generated using the BRASS SV calling pipeline were downloaded from the NCI GDC Data Portal for all patients in the MP2PRT cohort. We examined whether individual SVs were likely to have been formed via RAG recombination (*i.e.*, RAG-mediated), as previously described ^20^ and detailed in the Supplemental Methods. We assessed the per-patient burden and proportion of RAG-mediated SVs across molecular subtypes, and univariate and multivariable association tests were conducted to examine patient-level predictors of RAG-mediated traits (Supplemental Methods). A germline genetic dataset was generated from raw germline WGS data for 1,491 MP2PRT patients (Supplemental Methods). The cleaned single nucleotide polymorphism (SNP) genotype dataset was used to estimate genetic ancestry proportions in each patient, to calculate polygenic risk scores (PRS) for childhood ALL ^22^ and four lymphocyte-related blood cell traits previously associated with ALL risk ^23^, and to conduct multi-ancestry GWAS of RAG-mediated SV traits to examine the effects of known ALL risk SNPs and explore genome-wide associations (Supplemental Methods). Expression levels of both RAG1 and RAG2 were analyzed from RNA sequencing data in diagnostic samples from 1,465 of the MP2PRT patients (Supplemental Methods).

### Association between RAG-mediated SV traits and patient clinical outcome

We assessed the association of RAG recombination-mediated SVs with two binary clinical outcomes available in the Chang *et al*. dataset ^21^: relapse status (yes, no) and end of induction minimal residual disease (MRD) status (yes/no), two essential prognostic indicators for childhood ALL ^24^. First, we compared the frequency of off-target RAG-mediated SVs, overall and specific SV types, in patients who relapsed versus those who did not relapse using the Wilcoxon rank sum test for significance. We separately compared SVs with at least one RSS motif at a breakpoint, SVs with only one RSS at a breakpoint, and SVs with an RSS at both breakpoints between the relapse and non-relapse groups. To further investigate the association between off-target RAG-mediated SVs and relapse risk while accounting for relevant covariates, we conducted logistic regression models adjusted for age at leukemia diagnosis, sex, molecular subtype, and self-reported race/ethnicity. We also stratified the analysis by MRD status to examine whether RAG-mediated SVs may predict relapse risk independently of MRD. We repeated the regression models separately in the four molecular subgroups with the largest numbers of patients: *ETV6::RUNX1*, high hyperdiploid, *PAX5*alt, and B-other. Finally, we examined the association between off-target RAG-mediated SVs and MRD status overall and stratified by relapse status. Time-to-event outcomes were analyzed using weighted proportional hazards (PH) models (Supplemental Methods). Replication was performed using an independent cohort of T-cell ALL (T-ALL) patients ^25^ (Supplemental Methods).

## Results

### Landscape of RAG recombination-mediated SVs in B-ALL

Somatic SV calls were available for download in the GDC for n=1,496 patients with B-ALL included in the MP2PRT study, with patient-level information summarized in **Table S1** and characteristics of SVs in **Table 1**. Approximately 50% of deletions were located at Ig/TCR gene regions that are “on-target” for RAG recombination, whereas the majority of duplications (91.5%), inversions (68.5%), and translocations (97.3%) were “off-target”. The full-length RSS motif was identified in at least one breakpoint for 50.3% of SVs overall, 96.3% of on-target SVs, and 25.5% of off-target SVs, whereas 26.9% of overall, 67.6% of on-target, and 5.0% of off-target SVs harbored the RSS motif in both breakpoints (**Table 1**). Deletions were more likely to be RAG recombination-mediated than other SV types, particularly at off-target regions, in which ∼36% of deletions appeared to be RAG-mediated compared to <10% for other SV types (**Table 1, Figure 1**). Conversely, at Ig/TCR regions, a similar proportion of deletions (97.2%) and inversions (96.3%) and more than two-thirds of duplications and translocations were RAG-mediated.

**Figure 1.**
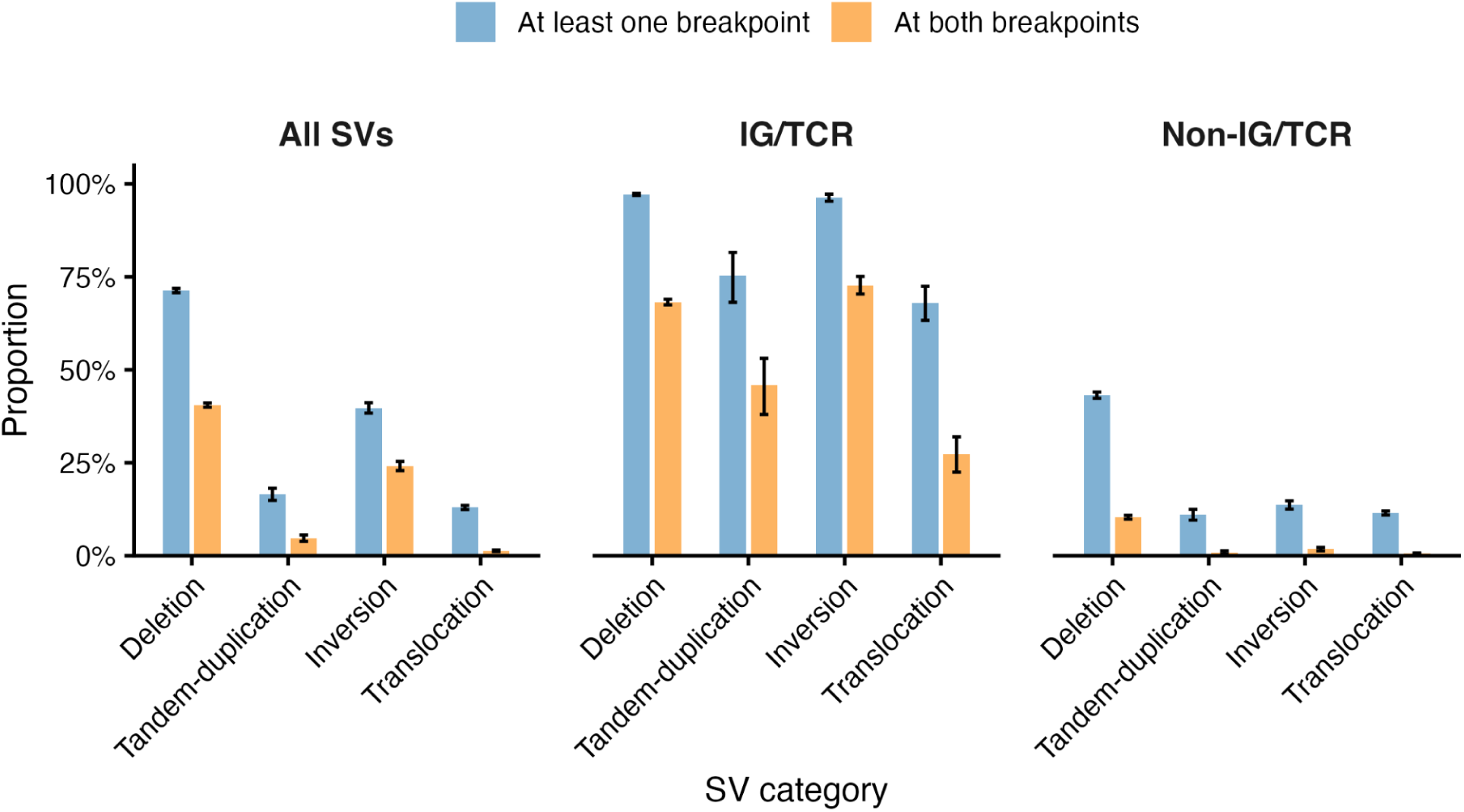
Proportion of RAG recombination-mediated structural variation by type and genomic region. Bar plots showing the proportion of structural variants (SVs) out of total SVs called in MP2PRT B-ALL patients in which the full-length RAG motif was detected in sequence surrounding at least one SV breakpoint or at both breakpoints. SVs are stratified by category (deletion, tandem-duplication, inversion, and translocation) and their genomic context (All SVs, Ig/TCR regions, non-Ig/TCR regions). SVs were more likely to be RAG-mediated at on-target Ig/TCR regions than in off-target non-Ig/TCR regions. At non-Ig/TCR regions, a higher proportion of deletions was RAG-mediated than for other SV types. Error bars represent 95% bootstrap confidence intervals.

Different SV types varied in size, with deletions, duplications, and inversions having median lengths of 61.5 Kb (range: 389bp-2,230,00Kb), 43.9 Kb (range: 819bp-223,000Kb), and 606.0 Kb (range: 33bp-218,000Kb), respectively, and the majority (94.0%) of deletions were smaller than 1 Mb in length (**Figure S2, Table S2**). On-target deletions were significantly larger than off-target ones (**Figure S3a**), whereas no significant difference was seen for overall SVs (**Figure S3b**). Limited to RAG-mediated events, there was no significant difference in size between on- and off-target deletions or SVs (**Figure S3C-D**).

### RAG recombination-mediated deletions at known driver genes

We examined the contribution of off-target RAG recombination to deletions overlapping known ALL driver genes. Among 12,769 deletions with gene annotations, we found evidence of RAG recombination for a similar proportion of deletions overlapping driver genes (42.6%, 2078/4873) and non-driver genes (43.5%, 3398/7806) (Chi-square p=0.336), suggesting that illegitimate RAG recombination occurs randomly at sites of RSS across the genome. *VPREB1* was the most frequently affected gene by RAG recombination, with deletions identified in 33.5% (499/1,491) of patients (**Figure 2, Table S3**). Although located in an immunoglobulin region on chromosome 22q11.22, *VPREB1* deletions have been reported to be independent of typical RAG recombination ^26^. In non-Ig/TCR regions, *CDKN2A*, *ETV6*, *PAX5*, and *CDKN2B* were the driver genes most frequently affected by off-target deletions. For other genes, including *IKZF1*, *RAG1*, *RAG2*, *CD200*, *BTLA*, *TBL1XR1*, *ADD3*, and in particular *SLX4IP*, a high proportion of overlapping deletions were RAG-mediated, and these deletions tended to be smaller in size than for genes in which the majority of overlapping deletions were not RAG-mediated (**Figure 2**). The bimodal size distribution of deletions overlapping *ETV6* and *ATF7IP*, both located on chromosome 12p, likely reflects the occurrence of focal deletions via RAG recombination in some patients versus loss of 12p through other mechanisms, *e.g.* nonhomologous end-joining^27^, in other patients.

**Figure 2.**
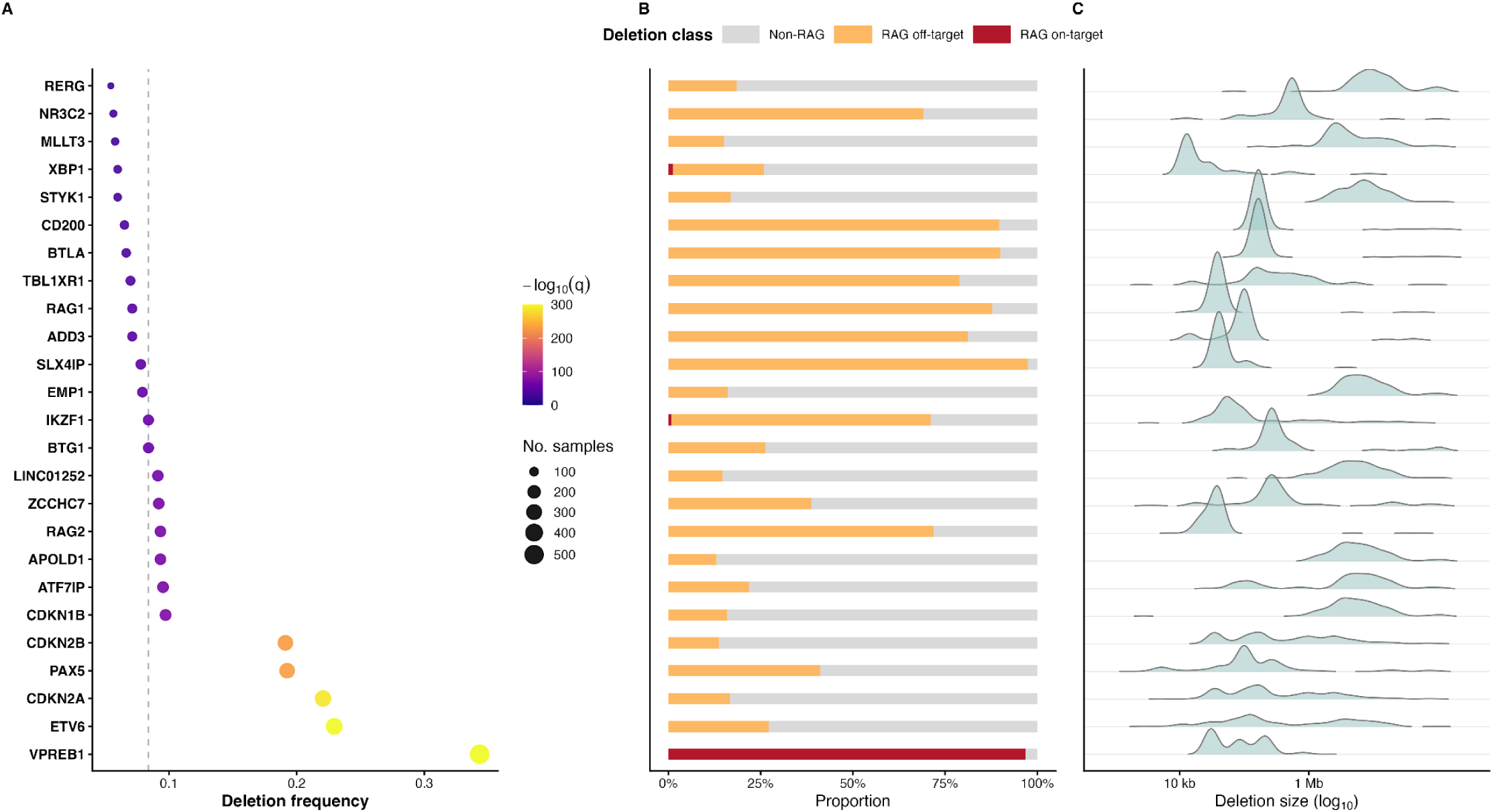
Characteristics of deletions overlapping childhood ALL driver genes. **A.** Deletion frequency of driver genes. Dot plot shows the frequency of patients with deletions overlapping known ALL driver genes across 1,491 B-ALL patients in MP2PRT with both structural variation and germline data. For each gene within each sample, only one deletion event was retained (the smallest deletion when multiple overlapped the same gene in the same sample.) Deletion frequency was calculated as the proportion of samples harboring at least one deletion affecting the gene. Point size represents the number of samples with deletions in that gene, and color indicates statistical significance (-log_10_(q-value), Benjamini-Hochberg adjusted). P-values were derived from a binomial test comparing the observed number of affected samples to a background deletion rate estimated across all driver genes. The dash vertical line denotes the median deletion frequency among the displayed genes. The 25 top-ranked genes by q values are shown. **B.** Deletion class composition across the top 25 driver genes. Stacked bar plots showing the proportion of deletions by mechanistic class, with the 25 top-ranked driver genes ordered as in panel A. Deletions were classified as non-RAG (gray), RAG off-target (orange; RAG-associated but outside V(D)J loci), or RAG on-target (red; RAG-associated within V(D)J loci). Bar length represents the proportion of deletions in each class per gene, with proportions calculated relative to the total number of samples harboring deletions in that gene. **C**. Distribution of deletion sizes across driver genes. Ridge density plots showing the distribution of deletion size for the same set of driver genes. For each gene-sample pair, a single deletion event was retained (the smallest deletion when multiple overlapped the same gene in the same sample). Deletion size is shown on a log_10_ scale. Density curves illustrate the relative distribution of deletion length per gene, enabling comparison of size patterns across recurrently deleted driver genes. Deletions of *ETV6* and *ATF7IP* demonstrate a bimodal size distribution, which likely corresponds to the occurrence of focal deletions via RAG recombination vs. loss of chromosome 12p arm through other mechanisms. Six genes have distributions peaking at the larger deletion size, including *RERG*, *STYK1*, *EMP1*, *LINC01252*, *APOLD1*, and *CDKN1B*; all are located on chromosome 12p, suggesting that these are co-deleted with *ETV6* through loss of chromosome 12p.

### RAG-mediated SVs across B-ALL molecular subtypes

Among 1,479 MP2PRT patients with B-ALL harboring off-target SVs and with available molecular subtype information, the frequency and proportion of RAG-mediated SVs varied substantially by subtype (**Figure 3**, **Table S4**). The *ETV6::RUNX1* subgroup had the highest burden of total SVs and RAG-mediated SVs as well as the highest per-patient mean number of off-target RAG-mediated SVs (mean: 8.14; 95% CI: 7.70, 8.56) among all subtypes (**Figure 3A-C**). The per-patient average number of off-target RAG-mediated SVs was also relatively high among *ETV6::RUNX1*-like (6.86; 95% CI: 5.33, 8.53) and *BCR::ABL1*-like (Ph-like) (7.14; 95% CI: 5.40, 9.20) patients. When considering the proportion among all off-target SVs in a subtype, there were similarly high proportions of RAG-mediated SVs among Ph-like (33.29%; 95%CI: 29.96%, 36.75%), *ETV6::RUNX1* (30.73%; 95%CI: 29.97%, 31.52%) and *DUX4*-rearranged (29.46%; 95%CI: 25.70%, 33.22%) subtypes (**Figure 3D**). Lower proportions of RAG-mediated SVs were observed in high hyperdiploid (17.68%; 95%CI: 16.78%, 18.60%) and *TCF3::PBX1* (10.42%; 95%CI: 7.90, 13.11%) subtypes. The pattern of results was similar for RAG-mediated SVs in which the RSS motif was detected at both breakpoints (**Figure S4**). Analysis of *RAG1* and *RAG2* gene expression data in MP2PRT B-ALL patients revealed that *RAG1* expression was highest in the subtypes with higher evidence of off-target RAG activity – *ETV6::RUNX1*, *ETV6::RUNX1*-like, and Ph-like subtypes – and generally lower in those with the lowest off-target RAG activity (**Figure 3E**). A distinct gene expression pattern was seen for *RAG2*, which did not appear to correlate with RAG-mediated SV frequency (**Figure 3F**).

**Figure 3.**
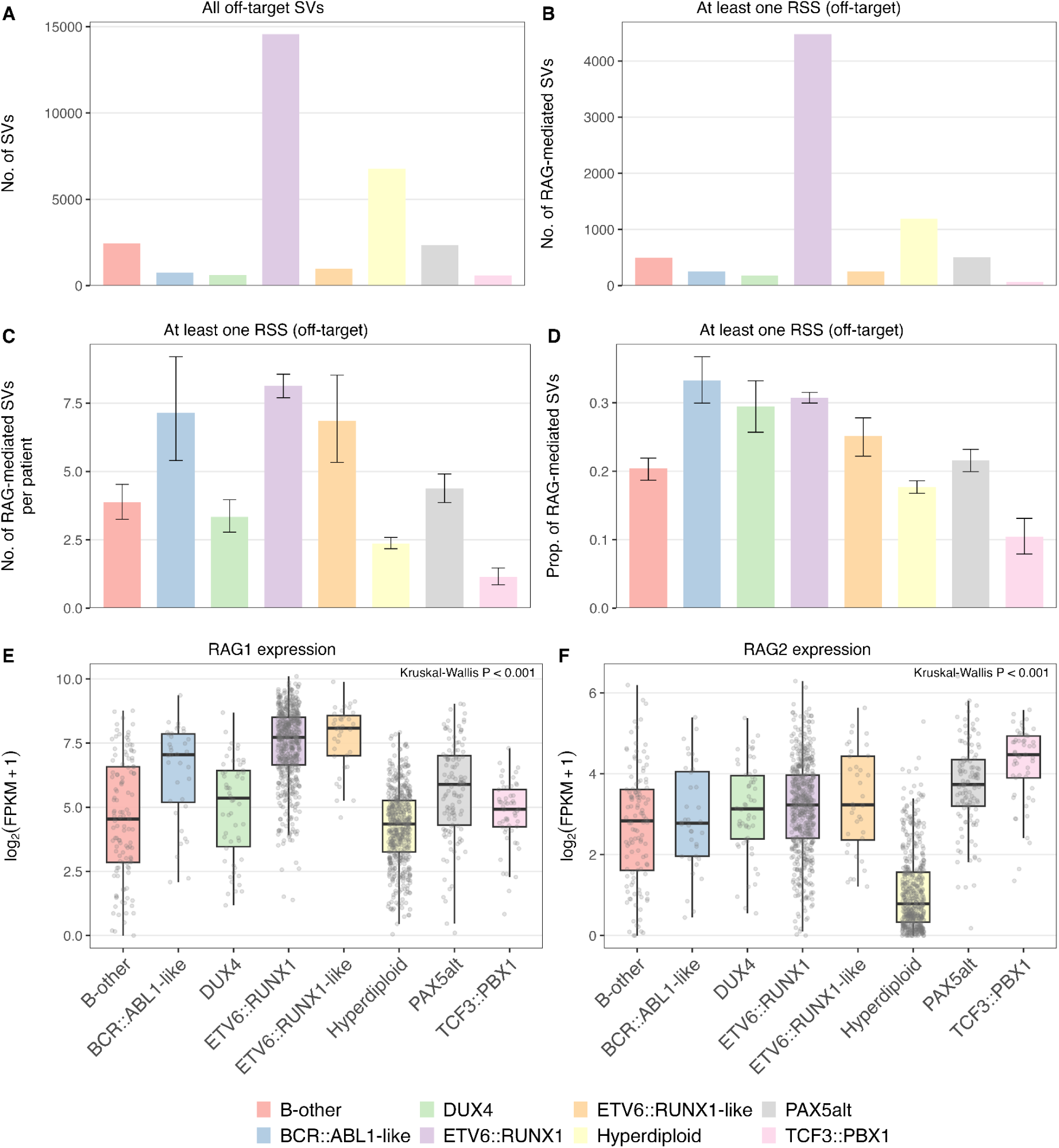
Number and proportion of off-target RAG-mediated structural variations (SVs) and RAG1/RAG2 expression by B-cell ALL molecular subtype. Bar plots showing: (**A**) the number of total off-target (non-Ig/TCR) SVs, (**B**) the number of total off-target RAG-mediated SVs (RSS at ≥1 breakpoint), (**C**) the average number of off-target RAG-mediated SVs per patient, and (**D**) the proportion of total off-target SVs that were RAG-mediated, across molecular subtypes of B-cell ALL in MP2PRT patients. Analyses were restricted to 1,479 patients with at least one off-target SV, including B-other (n=129), *BCR::ABL1*-like (n=35), *DUX4*-rearranged (n=54), *ETV6::RUNX1* (n=550), *ETV6::RUNX1*-like (n=36), Hyperdiploid (n=506), *PAX5*alt (n=115) and *TCF3::PBX1* (n=54) subtypes. Error bars represent 95% bootstrap confidence intervals. Panels (**E**) and (**F**) show *RAG1* and *RAG2* expression across all 1465 participants with tumor transcriptome profiling data, including B-other (n=117), *BCR::ABL1*-like (n=36), *DUX4*-rearranged (n=54), *ETV6::RUNX1* (n=552), *ETV6::RUNX1*-like (n=36), Hyperdiploid (n=501), *PAX5*alt (n=116) and *TCF3::PBX1* (n=53) subtypes. Gene expression levels calculated as fragments per kilobase of transcript per million mapped reads (FPKM) using RNA-seq data where forward and reverse strands are not distinguished.

### Patient-level predictors of RAG-mediated SVs

We next sought to understand whether individual-level patient characteristics may be associated with the prevalence of RAG-mediated SVs (see **Table S5** for modeling strategy). While patient sex showed no association, we found that older age-at-diagnosis was associated with a significantly higher number of RAG-mediated SVs, in particular for off-target SVs (**Figure S5**), and this remained significant in multivariable analyses (**Figure 4, Table S6-7**). While self-reported race/ethnicity did not show strong effects on RAG-mediated SV burden, Hispanic/Latino patients appeared to harbor a lower burden of on-target deletions compared with Non-Hispanic White patients. Similarly, increasing proportions of inferred IAM genetic ancestry were associated with reduced numbers of on-target RAG-mediated SVs (**Table S6**). Compared with *ETV6::RUNX1* B-ALL patients, most other subtypes were associated with lower burdens of total SVs and total, off-target, or on-target RAG-mediated SVs (**Figure 4, Table S6-7**). *ETV6::RUNX1*-like and Ph-like B-ALL patients showed no significant differences in SV traits compared to *ETV6::RUNX1* patients. While an increasing PRS for ALL was associated with a lower burden of RAG-mediated SVs in univariate analyses, these results were not significant in the multivariable model (**Figure 4**), suggesting that they may have been confounded by subtype. Lymphocyte-related trait PRS showed no associations with RAG-mediated SVs (**Figure 4**).

**Figure 4.**
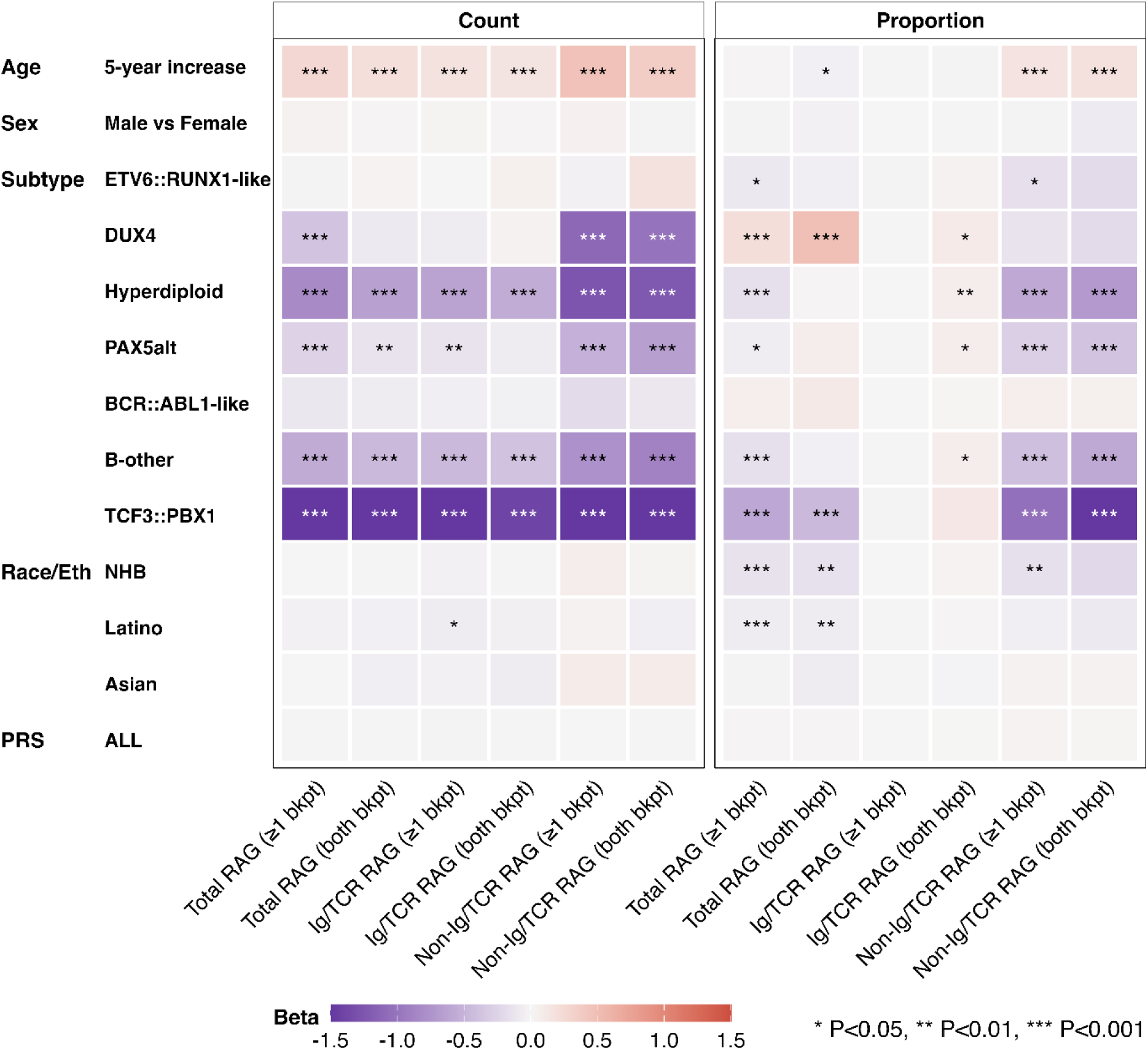
Patient-level characteristics and RAG-mediated structural variation traits. Multivariable association tests were conducted for SV traits, adjusted for age, sex, molecular subtype, self-reported race/ethnicity, and a polygenic risk score (PRS) for ALL. The heatmap displays results from negative binomial models for SV count burden and from negative binomial models with offset (total SVs) for SV proportion burden. For subtype analysis, *ETV6::RUNX1* subtype was the reference. For race/ethnicity (Race/Eth), non-Hispanic Whites were the reference, NHB = Non-Hispanic Black, Latino = Hispanic/Latino. * P<0.05, ** P<0.01, *** P<0.001.

As the number of RAG-mediated SVs may reflect the overall SV burden, we next examined the proportion of RAG-mediated SV traits as a function of all SVs detected within each individual. Associations between SV proportion traits and patient age-at-diagnosis and subtype were similar to those found for SV count traits (**Figure 4, Figure S5**). Patients diagnosed at an older age tended to have a greater proportion of off-target RAG-mediated SVs to total SVs, but not for on-target RAG-mediated SVs (**Table S8-9**). Compared with *ETV6::RUNX1*, most other subtypes exhibited a lower proportion of off-target RAG-mediated SVs but a higher proportion of on-target RAG-mediated SVs, with the strongest differences in high hyperdiploid and *TCF3::PBX1* B-ALL (**Figure 4**). Intriguingly, Non-Hispanic Black patients had significantly lower proportions of total and off-target RAG-mediated SVs than Non-Hispanic White patients, and proportions of AFR ancestry were associated with decreased proportions.

We next examined the association of 28 known childhood ALL risk loci^10, 22^ with RAG-mediated SV traits. SNPs at *ARID5B*, *CEBPE*, and *ERG* were associated with lower burdens of RAG-mediated SVs, while SNPs at 2q22.3 (near *RPL6P5*) and *IGF2BP1* were associated with both a higher burden and proportion of off-target RAG-mediated SVs (**Figure 5, Table S10**). The 2q22.3 and *IGF2BP1* SNPs were previously associated specifically with ETV6::RUNX1 B-ALL^28, 29^, and in analyses restricted to *ETV6::RUNX1* B-ALL patients, these loci were no longer significantly associated with RAG-mediated SVs (**Figure S6, Table S11**). Exploratory GWAS of RAG-mediated SV traits revealed several suggestive associations but none that reached genome-wide significance (**Figure S7-8)**. There was no evidence that germline variation at the *RAG1* or *RAG2* genes is associated with RAG-mediated SV traits (**Figure S9**).

**Figure 5.**
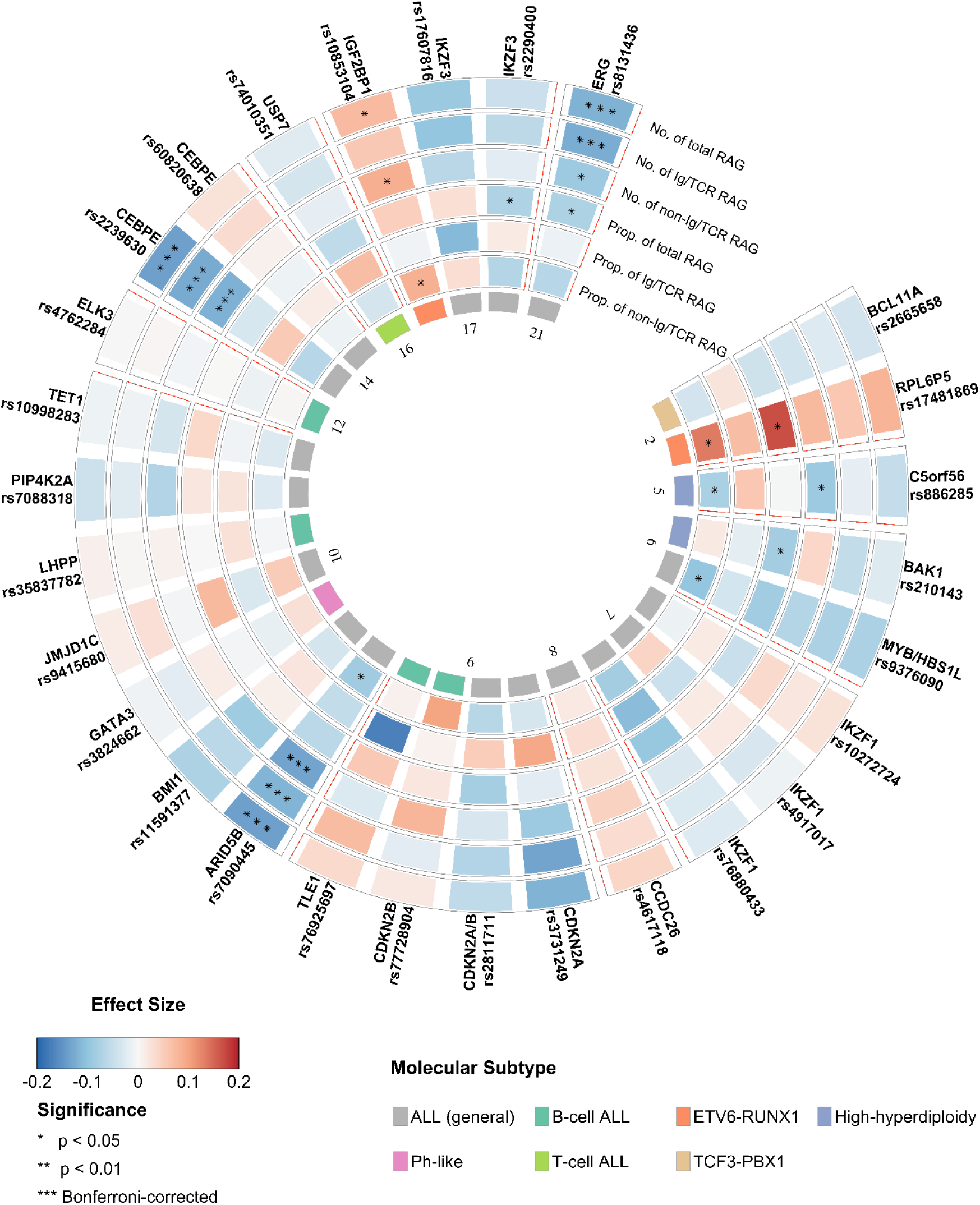
Childhood ALL GWAS risk loci and RAG-mediated structural variation traits. Associations between 28 reported risk loci from GWAS of childhood ALL and six RAG-mediated SV traits (number/proportion of total/on-target/off-target RAG-mediated SVs). Genes and single nucleotide polymorphism (SNP) rsIDs are depicted in the radials with one band per phenotype and divided by chromosomes. Colors indicate the magnitude and direction of the effect size. Gene–phenotype pairs reaching Bonferroni-corrected significance (*p* < 0.001786 (0.05/28)) are denoted by three asterisks.

### Association between RAG-mediated SVs and clinical outcome

In a recent analysis of diagnostic B-ALL samples from 150 patients, the frequency of SVs with RSS at one breakpoint was significantly higher among patients who subsequently relapsed (n=121) compared to non-relapsed patients (n=29)^18^. We leveraged available outcome data from MP2PRT and compared off-target RAG-mediated SVs in patients who subsequently relapsed (n=439) to those who did not (n=1,057). We found a significantly higher frequency of SVs with an RSS in at least one breakpoint (P=2.25 x 10^-3^) in diagnostic samples of patients who went on to relapse (**Figure 6A**). Limiting to SVs with an RSS at *only* one breakpoint revealed even greater significance with relapse (P=9.95 x 10^-4^). Examination of SV types revealed that off-target RAG-mediated duplications (P=0.039, RSS at only one breakpoint), inversions (P=0.018), and translocations (P=0.001) were significantly increased in the relapsed group compared to non-relapsed patients, whereas deletions were non-significantly increased (**Figure 6A**).

**Figure 6.**
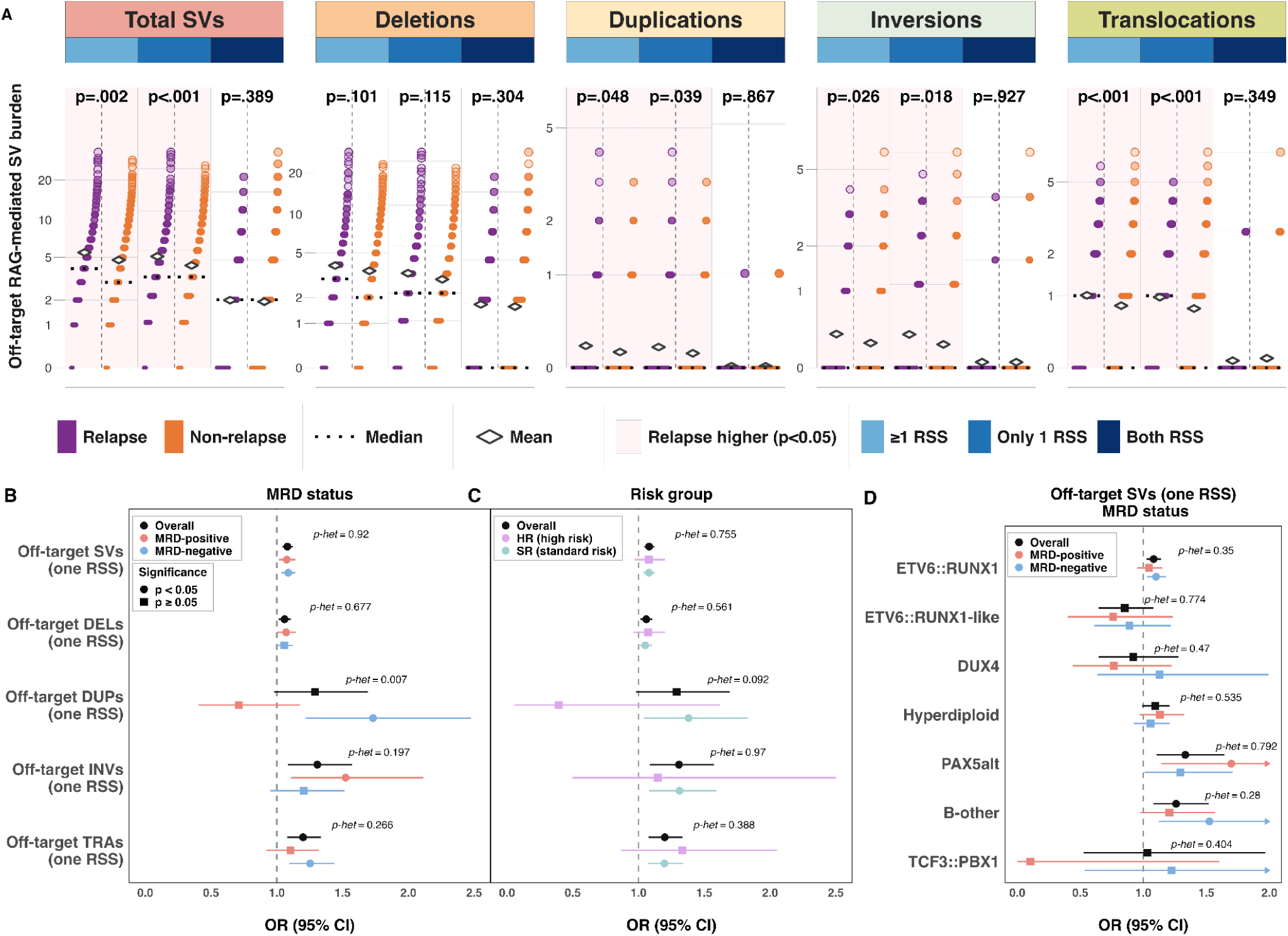
Off-target RAG-mediated structural variant burden in B-cell ALL patients by relapse status, SV type, and molecular subtype. **A**. Off-target SV burden per patient at diagnosis was compared between relapse and non-relapse patients across SV categories: off-target total SVs, deletions, duplications, inversions and translocations, each stratified by whether RSS was found in at least one breakpoint, at only one breakpoint, or at both breakpoints. The y-axis shows the number of off-target RAG-mediated SVs per patient, displayed on a ln(1 + x) scale to accommodate the large proportion of zero-count patients. Each dot represents one patient, ranked by SV count within each group. The dashed black horizontal line indicates the group median and diamonds indicate the mean. Purple shading indicates panels where the Relapse group has significantly higher burden (two-sided Wilcoxon rank-sum test, p < 0.05). **B**. Forest plot showing the association between off-target SVs with recombination signal sequence (RSS) at only one breakpoint at diagnosis and subsequent relapse status, stratified by minimal residual disease (MRD) status and SV category including deletions (Dels), duplications (Dups), inversions (INVs) and translocations (TRAs). Odds ratios (ORs) and 95% confidence intervals (CIs) are shown for different groups. P-values for heterogeneity (p-het) between different groups are displayed for each SV type. **C**. Forest plot showing the association between off-target SVs with RSS at only one breakpoint and relapse status, stratified by B-ALL patient risk group. **D**. Forest plot showing the association between off-target SVs with RSS at only one breakpoint and relapse status, stratified by MRD status and molecular subtype, including *ETV6::RUNX1*, *ETV6::RUNX1*-like, *DUX4*, hyperdiploid, *PAX5*alt, *BCR::ABL1*-like, B-other, and *TCF3::PBX1* subtypes.

We assessed the association between RAG-mediated SVs and both MRD status and relapse status while accounting for relevant covariates (age at diagnosis, sex, molecular subtype, race/ethnicity). First, we found that an increased frequency of RAG-mediated SVs with an RSS in at least one breakpoint, at both on- and off-target regions, was significantly associated with a higher odds of MRD positivity (**Table 2**). A higher proportion of RAG-mediated SVs at off-target regions, but not on-target regions, was strongly associated with MRD positivity (OR: 4.58 [1.98, 10.7]). Limiting analysis to SVs with an RSS at only one breakpoint revealed stronger effect sizes for the associations between RAG-mediated SVs and MRD status (**Table 2**).

In analysis of relapse, we observed that the total number of SVs with RSS at one breakpoint was strongly associated with relapse (OR: 1.08 [1.04, 1.12], P<.001) (**Table 2**). Importantly, effect sizes remained similar between patients who were MRD-negative (n=964) and MRD-positive (n=532) at end of induction (Phet=0.92). Among B-ALL patients who were MRD-negative, each additional SV with an RSS at only one breakpoint was associated with increased relapse risk with an OR of 1.09 (95% CI: 1.04-1.14, P<.001) (**Table 2**). We likewise observed that the total number of SVs – regardless of RAG-mediated status – was significantly associated with increased relapse risk (OR: 1.01 [1.01-1.02], P<.001). The number of off-target RAG-mediated SVs appeared to predominantly drive this observation, with an OR for relapse of 1.06 (95% CI: 1.01-1.11, P<.01) for each additional SV (**Table 2**). In sensitivity analysis limiting to SVs for which RSS were identified within 5 bp of breakpoint positions (Supplementary Methods), off-target RAG-mediated SVs remained significantly associated with relapse risk (**Table S12**).

Analysis of specific SV types revealed a similar trend of association between the burden of off-target RAG-mediated deletions and relapse risk in overall patients, while inversions, duplications, and translocations (*i.e.*, “non-deletion SVs”) with RSS at one breakpoint conferred even stronger effects with each additional SV associated with 1.31-, 1.29-, and 1.20-increased odds of relapse, respectively (**Figure 6B**, **Table S13**). Duplications showed a remarkably strong association with risk of relapse in MRD-negative patients (OR=1.73, 95% CI: 1.22, 2.48) but no association with relapse in MRD-positive patients (OR=0.71, 95% CI: 0.41, 1.18) (Phet=0.007), while effects for deletions, inversions, and translocations were similar between patient groups (Phet>0.1) (**Figure 6B-C**). Results were similar when calculated using weighted Cox regression tests (**Table S14**).

Analysis by molecular subgroup revealed that the frequency of off-target SVs with RSS at one breakpoint remained significantly associated with increased relapse risk across several genetic subtypes (**Figure 6D**). This association was observed in patients with more favorable subtypes *ETV6::RUNX1* (OR: 1.08 [1.03, 1.14]) and high hyperdiploidy (OR: 1.11 [1.00, 1.23]) as well as in higher risk subtypes *PAX5*alt (OR: 1.33 [1.10, 1.65]) and the B-other group (OR: 1.26 [1.08, 1.52]) (**Figure 6D**, **Table S15**). Similarly, results were consistent between patients classified with standard risk (n=1381) versus high-risk (n=115) disease (**Figure 6C**). *RAG1* and *RAG2* expression levels among patients who relapsed versus those who did not relapse displayed opposite patterns, with *RAG1* higher among non-relapsed and *RAG2* higher among relapsed patients (**Figure S10**). While this result was likely confounded by subtype (**Figure 3**), we did observe that both *RAG1* and *RAG2* expression were strongly positively associated with frequency of off-target RAG-mediated SVs (**Figure S11**).

We observed a generally increasing trend in relapse risk with increasing frequency of off-target RAG-mediated SVs (*p* for non-linearity = 0.020) (**Figure 7A-B, Figure S12**). A steep and statistically significant increased relapse risk was observed for ≥15 off-target RAG-mediated SVs, suggesting a potential threshold effect, and patients with ≥15 SVs (n=32) had a >3-fold increased risk of relapse (reference <median, HR: 3.23 [1.65,6.34]). Excluding deletions, a relatively large group of patients (n=229) harboring 3 or more off-target RAG-mediated SVs had a >1.8-fold increased hazard of relapse compared with patients with 0 non-deletion SVs (n=573) (HR: 1.88 [1.43, 3.49]) (**Figure 7C-D**). *ETV6::RUNX1* patients with ≥3 non-deletion SVs (n=126) had an almost 3-fold risk of relapse compared with patients with 0 non-deletion SVs (n=154) (HR: 2.85 [1.74, 4.66]), and a similar pattern was seen in MRD-negative *ETV6::RUNX1* patients (HR: 3.47 [1.86, 6.49]) (**Figure 8**).

**Figure 7.**
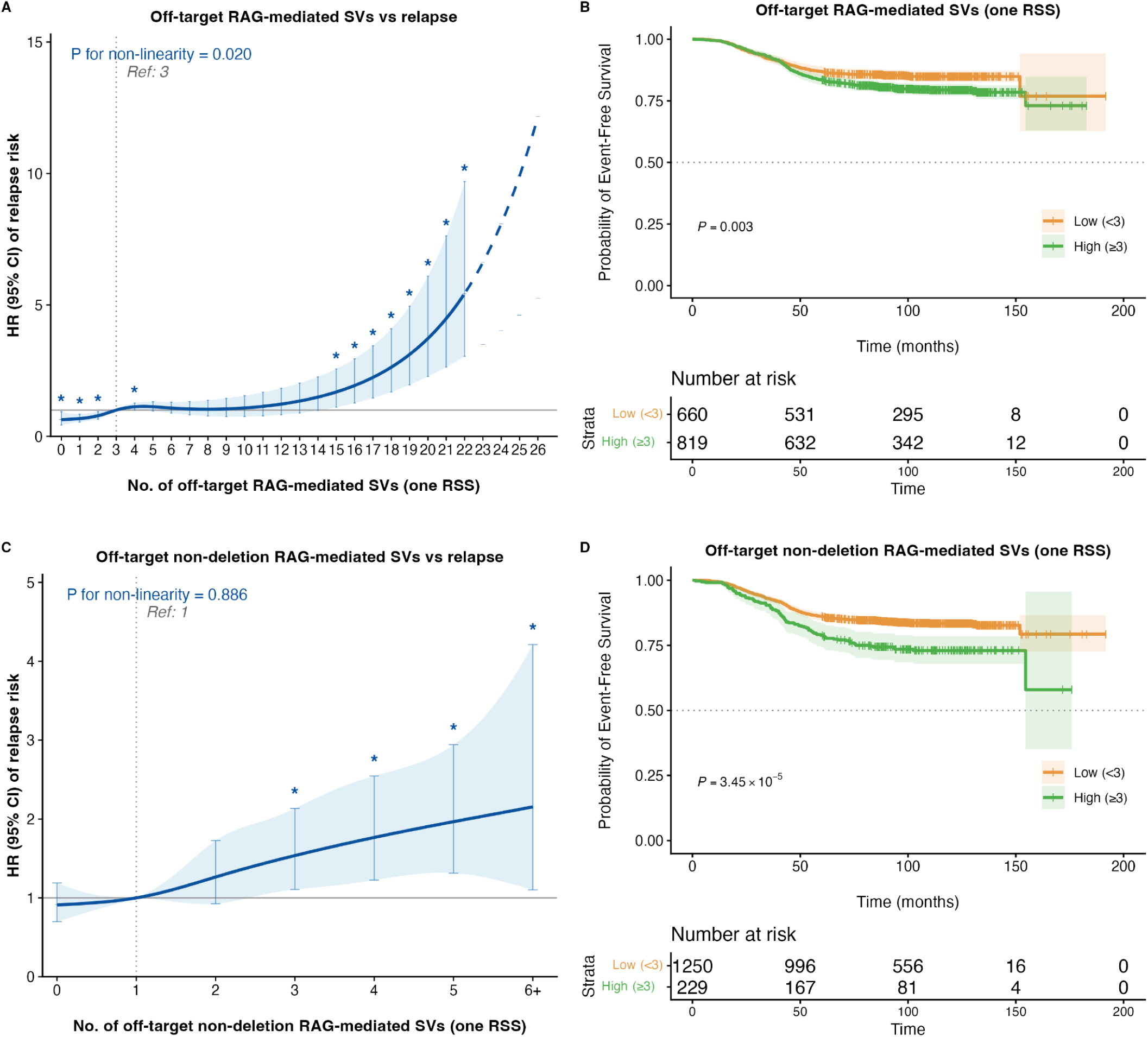
Off-target RAG-mediated structural variant burden and relapse risk across different SV count strata. **A**. Association between off-target RAG-mediated SVs with only one RSS and hazard ratio (HR) of relapse. Inverse-probability weighted (IPW) multivariable Cox proportional hazards model was fitted, adjusting for age, sex, race/ethnicity, molecular subtype and MRD status. The solid curve represents estimated HR relative to the cohort median SV count (reference), and the shaded region indicates the 95% confidence interval. The vertical dotted line and annotation mark the median reference value. Asterisks mark significant associations. P for non-linearity was assessed by the likelihood ratio test. **B**. IPW-weighted Kaplan-Meier curves for event-free survival (EFS) for relapse in patients stratified by SV burden: High (≥ median) versus Low (< median). **C**. Relapse risk and off-target RAG-mediated non-deletion SVs (duplications, inversions and translocations combined) with only one RSS. **D**. IPW-weighted Kaplan-Meier curves for EFS for relapse in patients stratified by off-target non-deletion RAG-mediated SVs.

**Figure 8.**
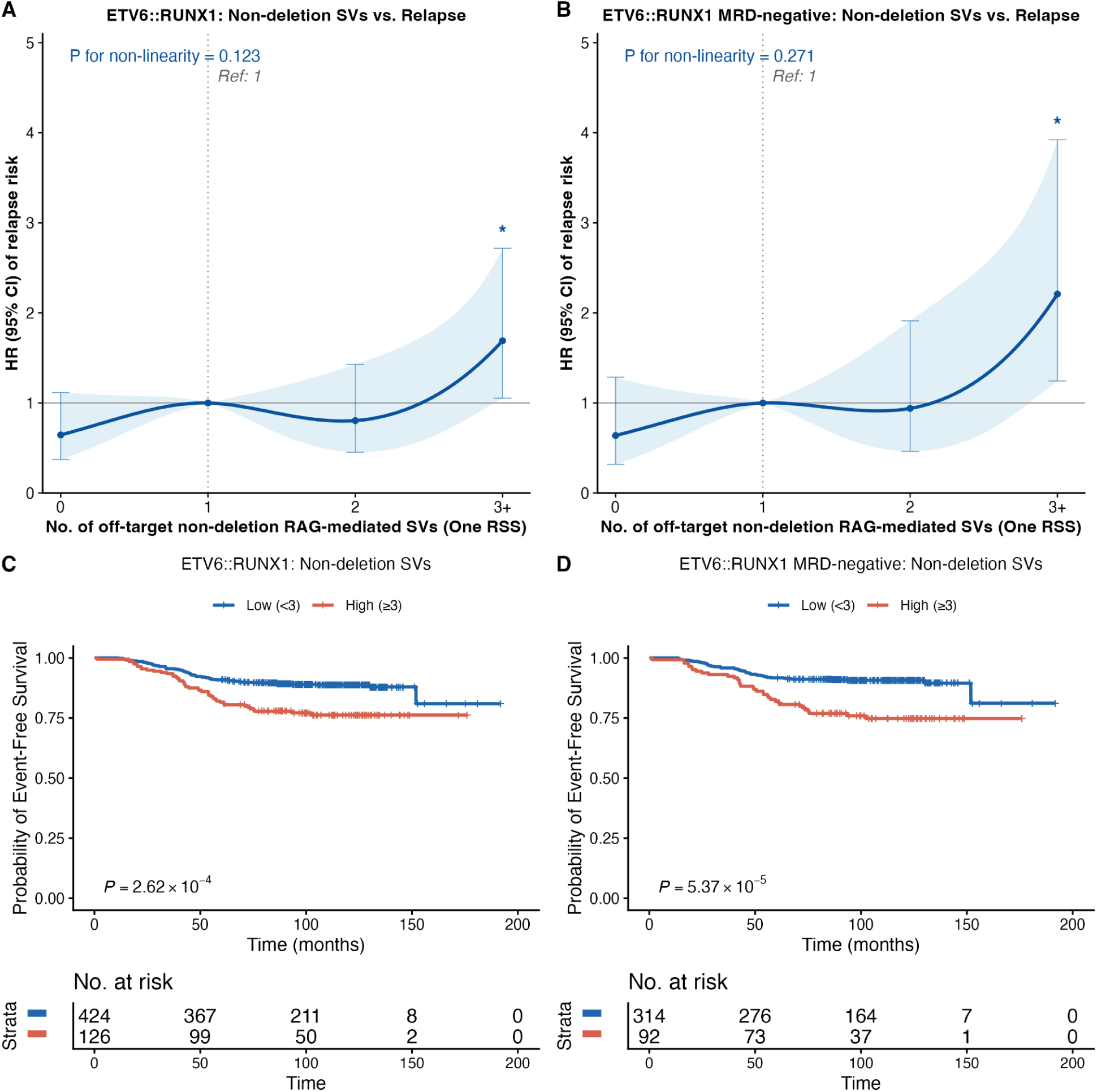
Off-target RAG-mediated non-deletion SVs and relapse-free survival in *ETV6::RUNX1* B-ALL patients. **A**. Association between off-target RAG-mediated non-deletion SVs with only one RSS and hazard ratio (HR) of relapse risk in *ETV6::RUNX1* patients. **B**. Off-target non-deletion SVs with only one RSS and relapse risk in MRD-negative *ETV6::RUNX1* patients **C**. Weighted Kaplan-Meier curves for event-free survival (EFS) for relapse in patients stratified by non-deletion SV burden (High ≥ median) versus low (< median) in *ETV6::RUNX1* patients. **D**. Weighted Kaplan-Meier curves for event-free survival (EFS) for relapse risk in MRD-negative *ETV6::RUNX1* patients.

### RAG-mediated SVs and clinical outcomes in T-ALL patients

Finally, we examined the association between RAG-mediated SVs at diagnosis and outcomes among 1,244 patients with T-ALL in COG trial AALL0434 ^25^, to assess whether RAG activity may impact outcomes across ALL immunophenotypes. While off-target RAG-mediated SVs with a single RSS were non-significantly associated with increased relapse risk in T-ALL (per SV, HR: 1.18, [0.99-1.39], p=0.0643), off-target RAG-mediated inversions (one RSS) showed significant association (HR: 1.47 [1.05-1.98], p=0.0259) (**Figure S13**, **Table S16**, **Supplemental Results**). T-ALL patients with ≥1 off-target RAG-mediated non-deletion SV had an ∼65% elevated risk of relapse (HR: 1.65, [1.11-2.46], p=0.016) compared with those without (**Figure S13**). Similar to B-ALL, associations were specific to RAG-mediated events.

## Discussion

In this study, we comprehensively characterize the landscape of RAG recombination–mediated somatic structural variation in patients with B-ALL, identify age-at-diagnosis and molecular subtype as predictors of off-target RAG-mediated SVs, and provide evidence that off-target RAG activity is an independent risk factor for disease relapse in B-ALL including in MRD-negative patients and across subtypes.

The MP2PRT study was designed to investigate genomic predictors of relapse risk in B-ALL patients ^21^ and identified deletions of specific genes at diagnosis, including *IKZF1*, *CDKN2A*, and *PAX5*, associated with subsequent relapse risk. Our data provide a potential molecular explanation for these findings by examining the total burden of deletions and SVs and the role of aberrant RAG recombination. We discovered that off-target RAG-mediated SVs at diagnosis are associated with an increased risk of B-ALL relapse. Significantly, we expanded upon the recently reported finding that off-target RAG-mediated SVs are higher in relapsed (n=121) versus non-relapsed (n=29) B-ALL patients ^18^. We observed the same association in a much larger patient cohort and additionally after adjusting for important predictors of relapse risk, including age-at-diagnosis and molecular subtype. Further, we assessed specific types of SVs and found that off-target RAG-mediated duplications, inversions, and translocations each appeared to confer a stronger risk of relapse than deletions, and that off-target RAG-mediated non-deletion SVs as a group were associated with a greater relapse risk than when including deletions. Importantly, the observed association between off-target RAG-mediated SVs and relapse risk was consistent across B-ALL molecular subgroups, with significant effects in the two most common subtypes, *ETV6::RUNX1* and high hyperdiploidy, and even stronger effects in the higher risk *PAX5*alt and B-other patient groups, and showed replication in T-ALL patients, suggesting that this phenomenon may be relevant for all patients agnostic of subtype.

Of additional important clinical importance, we discovered that having more off-target RAG-mediated SVs was equally associated with relapse in patients who were MRD-negative compared to those who were MRD-positive at end of induction. This suggests that the burden of off-target RAG recombination may serve as an independent risk factor for relapse that offers novel prognostic information even beyond MRD status, a critically important variable for dictating post-induction treatment intensity ^30^. Crucially, we identified a group of n=92 *ETV6::RUNX1* patients who harbored at least 3 off-target RAG-mediated *non-deletion* SVs at diagnosis and, despite being MRD-negative at end of induction, had an almost 4-fold risk of relapse. Pending replication of these findings in the context of the high overall cure rates achieved with modern chemo-immunotherapy protocols ^1^, patients with high numbers of RAG-mediated SVs may ultimately benefit from future trial initiatives to investigate risk-directed treatment intensification.

The increased numbers of off-target RAG-mediated SVs in patients with B-ALL who experienced relapse may result from ongoing RAG activity or high clonal complexity that promote treatment resistance ^18^. This is supported by the strong significant association we observed between RAG-mediated SVs and MRD positivity. Further research is required to understand how this may confer an increased survival capacity of leukemia subclones under treatment. Whether the burden of off-target RAG-mediated SVs at diagnosis correlates with pretreatment Ig clonal composition, as measured by IgH high-throughput sequencing (HTS) for sensitive MRD detection, will also be important to investigate given context-dependent associations between pre-treatment IgH rearrangement state and outcome in patients with B-ALL ^31, 32^. Future studies are also required to discern which biomarkers of RAG activity, *e.g.*, levels of ESCs, RAG gene expression, or RAG-mediated SVs or specific SV types, may be most clinically important for refining B-ALL risk stratification. In addition, it will be important to study whether increased off-target RAG recombination confers risk of relapse for B-ALL patients in the advent of novel immunotherapies, such as blinatumomab^1^. Finally, while we uncovered that off-target RAG-mediated SVs may also contribute to patient outcomes in T-ALL, in which illegitimate RAG recombination is an important driver of somatic alterations as well ^25, 33^, further studies in cohorts with larger numbers of relapsed patients will be required to confirm a role for off-target RAG activity in outcomes across ALL immunophenotypes.

Our findings confirm illegitimate off-target RAG recombination as a predominant driver of somatic SV formation in B-ALL leukemogenesis, in particular for the generation of second-hit deletions. Among known ALL driver genes, *VPREB1* emerged as the most frequently deleted by RAG recombination, consistent with its encoding a component of the surrogate light chain of the pre-B cell receptor and location within the immunoglobulin lambda locus (IGL), a position inherently susceptible to RAG recombination activity ^26^. The vast majority of *VPREB1* deletions observed in childhood B-ALL are focal and do not extend to the lambda light chain VDJ junction, supporting that these deletions do not occur via on-target RAG recombination ^26^. Deletions overlapping other driver genes, including *IKZF1*, *RAG1*, *RAG2*, *CD200*, *BTLA*, *TBL1XR1*, *ADD3*, and *SLX4IP*, were largely RAG-mediated, likely due to the proximity of RSS to these genes. In particular, focal deletions at *SLX4IP* were almost always found to be RAG-mediated, as previously reported in a European cohort of ALL patients ^34^.

We confirm RAG-mediated recombination as a major mutational driver in *ETV6::RUNX1* fusion B-ALL ^12^, and in other subtypes in which RAG-mediated SVs have not been comprehensively studied, including *DUX4*-rearranged ^35^, Ph-like, and ETV6::RUNX1-like B-ALL. Gene expression analysis in B-ALL patients at diagnosis suggested that variation in expression of *RAG1*, but not *RAG2*, may drive these subtype differences in SV formation. The *ETV6::RUNX1* fusion has been shown to bind to regulatory regions of *RAG1* resulting in overexpression and likely increased aberrant RAG activity ^13, 36^. Moreover, as expression of *RAG1* and *RAG2* varies across different stages of B-lymphocyte development, the preleukemic cell of origin likely influences RAG activity across B-ALL subtypes. While *ETV6::RUNX1* has been proposed to originate in CD34+CD19+ precursor B-cells, *TCF3::PBX1* fusions are thought to arise at a later stage of development ^37–40^, which may at least in part explain differences in *RAG* expression and in the burden of RAG-mediated SVs seen between these two subtypes.

Patient age-at-diagnosis was also a strong determinant of RAG-mediated SVs in this study, with an older age-at-diagnosis positively associated with both the burden and proportion of RAG-mediated SVs, in particular for off-target RAG-mediated SVs with RSS at one breakpoint. While a positive association between patient age and number of SVs may reflect an accumulation of RAG-mediated events over time, further research is required to assess whether the burden of ESCs increases in B-ALL patients with increasing age-at-diagnosis and continues to contribute to aberrant RAG activity. In addition, while differences in RAG-mediated traits are unlikely to contribute to the increased B-ALL risk in males or in children of Hispanic/Latino ethnicity based on our observations, further research is warranted to examine whether the association between Non-Hispanic Black ethnicity and lower proportions of off-target RAG-mediated SVs may contribute to the lower incidence of B-ALL in this population ^41^. The lack of a strong association between germline genetic variants and RAG-mediated SV traits suggests that, beyond B-ALL subtype and patient age, environmental factors such as early-life infections ^6^ and tobacco smoke exposure ^20^ may play a greater role in determining the extent of RAG activity in leukemic cells. We note that our GWAS was limited in sample size and consisted of case-only analyses. Unmeasured factors that may influence both RAG activity and ALL risk, such as frequent childhood infections, could conceal the effects of genetic variation among patients. Further studies are warranted to investigate potential germline genetic effects on RAG recombination in case-control studies of ALL, or among healthy individuals via single-cell sequencing approaches ^42^.

This is, to our knowledge, the most comprehensive assessment of aberrant RAG recombination-mediated SVs in B-ALL patients. Our strategy to identify putatively RAG-mediated SVs is supported by the expected frequencies and proportions of events that we observed at Ig/TCR vs. off-target genomic regions and across different SV types, as well as the enrichment of RAG-mediated deletions in *ETV6::RUNX1* B-ALL patients ^12^ and in the *ETV6::RUNX1*-like subtype ^43^. Our study has some limitations. The B-ALL patient cohort was enriched for relapsed cases due to the MP2PRT study design ^21^, thus ascertainment bias should be considered when interpreting the generalizability of our findings, although this is unlikely to confound the association between RAG-mediated SVs and clinical outcomes. The MP2PRT cohort was also selected mostly for standard risk B-ALL patients, and thus several COG-defined unfavorable subtypes such as *KMT2A*-rearranged, hypodiploid, and iAMP21, were missing or underrepresented in our analyses. Another limitation was the absence of epidemiological data on environmental factors, such as early-life tobacco exposure ^20^ and cytomegalovirus (CMV) infection ^7, 44^, two potential determinants for RAG-mediated recombination activity.

In summary, our findings implicate RAG-mediated genetic instability as a key driver of leukemogenic structural variation in pediatric B-ALL and introduce off-target RAG-mediated SV burden as an independent predictor of relapse risk, irrespective of MRD status. These data underscore the clinical relevance of off-target RAG-mediated SV accumulation as a previously underappreciated prognostic indicator while also providing mechanistic insight into the role of aberrant RAG activity in leukemia pathogenesis.

## Supporting information

Main Tables

## Data Availability

All data produced in the present study are available upon reasonable request to the authors.

## Acknowledgements

We thank Drs. Karen Rabin, Kara Davis, and Saro Armenian for valuable discussions and suggestions. This work was supported by grants from the National Institutes of Health (NIH, R01CA292941 to A.J.d.S. and V.G.S., and R01CA262263 to A.J.d.S., C.W.K.C., J.L.W.). S. S. is a Scholar of the St. Baldrick’s Foundation and is supported by grants from the Norman and Sadie Lee Foundation and the Dake-Wilson Family Pediatric Accelerator Fund. A.J.d.S. is a Scholar of Blood Cancer United. The content and conclusions from this work do not reflect the official views of the sponsors. Computation of this work was supported by USC’s Center for Advanced Research Computing (https://www.carc.usc.edu/).

This study makes use of data from the Molecular Profiling to Predict Responses to Therapy (MP2PRT) study available from dbGaP (accession number: phs002005.v1.p1). The MP2PRT study was supported by NIH grants U10CA98543, U10CA98413, U10CA180886, and U10CA180899.

