## Supplementary Methods for "RAG-mediated structural variation and its impact on relapse risk in acute lymphoblastic leukemia"

**Supplemental Methods**

**Study subjects**

This study used data from the Molecular Profiling to Predict Responses to Therapy (MP2PRT) study of B-ALL (dbGaP accession number phs002005.v1.p1), which included 1,496 patients enrolled in Children’s Oncology Group (COG) trials ^1^. We assigned patients to race/ethnicity groups as follows: patients reported as "Hispanic or Latino" ethnicity regardless of race were grouped as "Hispanic/Latino" (n=327), "White" race and not Hispanic or Latino were grouped as "Non-Hispanic Whites" (n=907), "Black or African American" and not Hispanic or Latino were grouped as "Non-Hispanic Blacks" (n=79), "Asian" and not Hispanic or Latino were grouped as Asian (n=52), "American Indian or Alaska Native" (AIAN) (n=6), "Native Hawaiian Pacific Islander" (n=2), and unknown ethnicity and/or unknown race if not reported as Hispanic or Latino were grouped as "Unknown" (n=123). We limited our molecular subgroup analyses to subtypes present in at least 2% of the patient cohort, including: *ETV6::RUNX1* (n=557), high hyperdiploid (n=514), *PAX5*alt (n=116), *TCF3::PBX1* (n=54), *DUX4*-rearranged (n=54), *ETV6::RUNX1*-like (n=36), and *BCR::ABL1*-like (or Ph-like, n=36). Patients classified as “B-other” (n=44) were grouped with all remaining subtypes as B-other (total n=129), including: *ZNF384*-rearranged (n=25), Down syndrome B-ALL (n=19), *NUTM1* (n=8), *MEF2D* (n=7), iAMP21 (n=5), *IKZF1* N159Y (n=5), *PAX5* P80R (n=4), *TCF3/4::HLF* (n=4), *BCL2/MYC* (n=3), *KMT2A* (n=2), *ZNF384*-like (n=1), *ZEB2/CEBPE* (n=1), *TCF3::PBX1*-like (n=1).

### **Structural variant breakpoint analysis**

We examined whether individual SVs were likely to have been formed via V(D)J recombination, or RAG recombination, as previously described ^2^. In brief, we investigated the presence of full-length recombination signal sequence (RSS) motifs in the +/-50bp sequence flanking each SV breakpoint using the Find Individual Motif Occurrences (FIMO) tool in MEME suite v5.5.5 (*P*<10-4) ^3, 4^. Cryptic RSS sites were identified using the position-specific scoring matrix which accommodates sequence variation within the motif ^5, 6^ assuming the background rate of 0.2 for C/G and 0.3 for A/T. The full-length RSSs are conserved DNA motifs that guide RAG1/RAG2 proteins during V(D)J recombination in lymphocyte development, consisting of a heptamer (CACAGTG) and a nonamer (ACAAAAACC) separated by a 12 or 23 base pair spacer. SVs were then annotated according to whether an RSS motif was identified at one or both breakpoints, with presence of the full-length RSS motif in the flanking sequence of at least one breakpoint considered as strong evidence of RAG recombination. SVs were further annotated based on whether at least one breakpoint overlapped with immunoglobulin/T-cell receptor (Ig/TCR) gene regions (*i.e.*, IgH, IgK, IgL, TRB, TRA/TRD, TRG) or whether both breakpoints were outside of +/-1000 bp of Ig/TCR regions (non-Ig/TCR). SVs that overlapped either Ig/TCR regions or non-Ig/TCR regions and had RSS motifs in at least one breakpoint sequence were denoted to be formed via “on-target” or “off-target” RAG recombination, respectively. We assessed the total number and proportion of the different types of SV (deletions, tandem duplications, inversions, translocations) in the BRASS results that were identified as RAG recombination-mediated (one or both breakpoints), and at on-target versus off-target regions. We also assessed the overlap of deletions with known ALL driver genes based on prior studies ^7–10^ using AnnotSV3.5 ^11^. For each gene within each sample, only one deletion event was retained (for overlapping deletions within the same gene and sample, the smallest deletion was selected). Deletion frequency was calculated as the proportion of samples harboring at least one deletion overlapping the gene.

Our overall analysis of RAG recombination-mediated SVs included a lenient threshold of +/-50 bp distance of the RSS from the breakpoint positions due to the uncertainty of precise breakpoint locations, for instance due to the presence of repetitive DNA sequence spanning breakpoints. Therefore, to support the robustness of our findings, we performed sensitivity analyses in which we limited the selection of RAG-mediated SVs to those in which full length RSS motifs beginning with CAC or GTG were identified to start within a range of distances from the breakpoints, including from only +/-5 bp, +/-10 bp, and +/-20 bp, and then we repeated the association testing between off-target RAG-mediated SVs and patient outcomes.

**Germline genotype data generation**

A germline genetic dataset was generated from raw germline WGS data for the 1491 MP2PRT patients with B-ALL in the NCI GDC Data Portal. First, we extracted a set of 1,369,675 single nucleotide polymorphism (SNP) genotypes equivalent to those included on the Illumina Global Diversity Array (GDA) by using SAMtools ^12^ to filter patient germline sample BAM files for reads that overlapped autosomal markers on the GDA manifest file. The genotype data were then processed using the Genome Analysis Toolkit (GATK v4.6.1.0) Best Practices workflow for germline short variant discovery ^13^. During pre-imputation QC, variants were excluded if they had low call rate (<95%), low minor allele frequency (MAF, <0.01), violation of Hardy-Weinberg equilibrium (p<10^-7^), and missingness differences by sex (p<10^-7^). Individuals were removed if they had a call rate lower than 10%. Imputation was then performed using the TOPMED Imputation Server ^14–16^. Post-imputation, individual genotypes with a posterior probability ≥ 0.85 were retained, and we removed variants with imputation quality R^2^ <0.5, missingness greater than 0.05 and MAF less than 0.01, deviations from Hardy-Weinberg equilibrium (p<10^-7^), and missingness differences by sex (p<10^-7^). Following post-imputation QC, a total of 8,202,037 SNPs were retained for downstream analyses.

**Estimation of genetic ancestry**

Global genetic ancestry was inferred from the cleaned SNP genotype data using RFMix (v.2.03)^17^. The model was trained on a selection of 1,000 Genomes Phase 3 Reference Panel data representing five superpopulations - African (n=883), Admixed American (n=535), East Asian (n=621), European (n=562), and South Asian (n=661) - to infer local ancestry estimates at windows across the genome for each individual. We used a default window size of 0.2 cM and assumed eight generations since admixture to optimize estimates for African and European admixture. In each patient, the proportions of each chromosome assigned to each ancestry group were summed to calculate global ancestry proportions of European (EUR), African (AFR), Indigenous American (IAM), East Asian (EAS) and South Asian (SAS).

**Polygenic risk score calculation**

To explore the role of common genetic variation associated with ALL risk in the formation of RAG-mediated SVs, we calculated a polygenic risk score (PRS) including SNPs identified in prior GWAS of childhood ALL ^18^^,^^19^, with SNP weights calculated as the log odds ratio of the ALL association. We also examined PRS for four lymphocyte-related blood cell traits that were previously associated with childhood ALL risk: lymphocyte counts, lymphocyte-to-monocyte ratio (LMR), neutrophil-to-lymphocyte ratio (NLR), and platelet-to-lymphocyte ratio (PLR) ^19^. Blood cell trait PRS included genetic instruments selected from our prior multi-trait analysis of GWAS (MTAG), which reflect independent effects on the different blood cell traits, with SNPs weighted by beta coefficients for the relevant trait. PRS were standardized within the study sample by subtracting the mean PRS and dividing by the standard deviation, yielding Z-scored PRS with mean 0 and unit variance.

### **Association between patient characteristics and RAG-mediated SV traits**

SV count traits were defined as per-person counts of SVs overall (total) or separately in on-target (Ig/TCR) or off-target (non-Ig/TCR) regions. A negative binomial model was fitted for the total number of RAG-mediated SVs and the number of on-target RAG-mediated SVs. SV proportion (fraction) traits were defined as the per-patient fraction of the total, on-target or off-target SVs that were RAG-mediated in that patient. For each SV proportion trait, we fitted negative binomial models with an offset term to model the RAG-mediated SV rate relative to the overall SV burden in each patient. Samples with zero total SVs (*i.e.,* denominator equal to zero) were excluded because the offset was undefined. Additionally, the beta-binomial model was fitted as an orthogonal approach for modeling the proportions data. In the univariate association tests, we included age at leukemia diagnosis, sex (female as reference), molecular subtype (*ETV6*::*RUNX1* [reference], *ETV6*::*RUNX1*-like, *DUX4*, Hyperdiploid, *PAX5*alt, *BCR::ABL1*-like, B-other), self-reported race/ethnicity (non-Hispanic White [reference], non-Hispanic Black, Hispanic/Latino, Asian), estimated global ancestry proportions (EUR [reference], AFR, AMR, EAS, SAS), standardized ALL PRS as well as four standardized lymphocyte-related blood cell traits PRS (*i.e.*, lymphocyte counts, LMR, NLR, and PLR) as our *a priori* independent predictors for RAG-mediated SV traits. In the multivariable model, based on prior knowledge and univariate selection we included age at diagnosis, sex, molecular subtype, and self-reported race/ethnicity and the standardized ALL PRS.

### **Genome-wide association studies of RAG-mediated SV traits**

Using the post-imputation SNP genotype data, we conducted multi-ancestry GWAS of several RAG-mediated SV traits using the GENetic EStimation and Inference in Structured samples (GENESIS) workflow in *R* ^20^. GENESIS is well suited for WGS data in a multi-ancestry study as it appropriately accounts for cryptic sample relatedness and population structure at a genome-wide scale. The six phenotypes included in the GWAS were the number or proportion of total, on-target, or off-target RAG-mediated SVs. To ensure normality and minimize the influence of outliers, each phenotype was transformed using a rank-based inverse normalization method. We performed a standard linear mixed model (LMM) GWAS. The first model was fit under the null hypothesis that each SNP has no effect, adjusting for sex, age at leukemia diagnosis, and the first 10 genetic ancestry-related principal components (PCs) generated using PC-AiR ^21^ and PC-Relate ^22^. We included the 4th-degree sparse empirical kinship matrix (KM) computed with PC-Relate ^22^ as a random effect to account for genetic relatedness. The residuals from this null model were subsequently used to perform single-variant genome-wide score tests. For the GWAS, we used a P-value threshold of 5 x 10^-8^ to adjust for the number of independent variants (but did not adjust for the number of traits, which were not truly independent). In addition, we examined the association of a list of established childhood ALL risk SNPs (n=28) ^23^, using a significance threshold of P=0.0018 (0.05/28). We also explored whether SNPs across the *RAG1* and *RAG2* gene region at chromosome 11p12-p13 showed evidence of association with RAG-mediated SV traits, using a lenient *P*-value threshold of 1.45 x 10^-5^ based on n=3433 SNPs (P < 0.05/3433) in a +/- 500kb window centered on the *RAG1*/*RAG2* genes. Finally, we performed a sensitivity analysis among B-ALL patients with the *ETV6*::*RUNX1* subtype (n=554) to examine the effects of known ALL risk SNPs. Based on PC-Relate results, the level of relatedness among *ETV6*::*RUNX1* patients was minimal, thus we conducted a standard GWAS using linear regression in PLINK2, adjusting for age at diagnosis, sex, and the top three PCs (due to smaller sample size compared with the full GWAS dataset).

**RAG1/RAG2 gene expression analysis**

Expression levels of RAG1 and RAG2 genes were obtained from RNA sequencing data in diagnostic samples from 1,465 of the MP2PRT B-ALL patients, including n=1,288 with whole bone marrow and n=177 with peripheral whole blood specimens. The derived Fragments Per Kilobase of transcript per Million mapped reads (FPKM) data were extracted from the GDC portal. RNA-sequencing and data pre-processing was described previously ^1^. Kruskal-Wallis tests were used to assess overall differences in RAG1/RAG2 gene expression by molecular subgroup, and Wilcoxon rank-sum tests were used to evaluate gene expression differences by relapse status. All tests were two-sided and p <0.05 was considered statistically significant. We further examined the association between RAG1/RAG2 gene expression and frequency of off-target SV events. Spearman’s correlation coefficient was used to assess the association between RAG1/RA2 log2 -transformed FPKM unstranded counts (plus pseudocount) and number of off-target SVs with RSS in at least one breakpoint, at only one breakpoint, or at both breakpoints.

**Differential expression and gene set enrichment analysis**

To determine whether differential expression results were driven by RAG-mediated SVs, we used DESeq2 (v.1.50.2) ^24^. We filtered out low count genes ($\leq$10) and applied VST transformation. The known variable age and molecular subtype were adjusted for the count matrix. Genes with |log2 fold change| > 1 and adjusted p-value (Benjamini-Hochberg) < 0.05 were considered differentially expressed. Gene set enrichment analysis (GSEA) was performed with gene sets obtained from MSigDB v2023.1, including Hallmark, KEGG, Gene Ontology and Reactome collections ^25, 26^. Genes were pre-classified by off-target RAG-mediated SVs, on-target RAG-mediated SVs vs non-RAG-mediated SVs. Statistical significance was defined as normalized enrichment score (NES) |NES| >1.5 and FDR q-value <0.05 (1,000 permutations, minimum gene set size=15, maximum =500).

**Survival analysis**

A total of 1,479 MP2PRT B-ALL patients had both genomic and survival data available. Multivariable weighted Cox proportional hazard (PH) model with time on the study as the timescale was used to estimate the hazard ratios (HRs) and 95% confidence intervals (CI) of time-to-event outcomes including event-free survival (EFS, relapse vs. none), disease-free survival, and overall survival, adjusting for age at diagnosis, sex, self-reported race/ethnicity, molecular subtype, and MRD status. Standard tests using Schoenfeld residuals suggested no evidence of violation of proportional hazards assumptions. To account for relapse enrichment sampling in the MP2PRT study, inverse probability weights (IPW) were derived from the estimated sampling rate as 0.442 for cases (relapse event) and 0.029 for controls (none), as described by Chang et al. ^1^. Weighted Kaplan Meier curves were constructed for two comparison groups (median as the cutoff) within the IPW-weighted pseudo-population to visualize the probability of EFS over time. P-values were derived from the multivariable weighted Cox regression models adjusting for the same set of covariates as appropriate. Weighted Fine-gray competing risk models were fitted to estimate the cumulative risk of relapse in the presence of non-relapse mortality as the competing event.

**External replication in T-cell ALL patients**

A total of 1,244 T-cell ALL (T-ALL) patients from COG clinical trial AALL0434 ^27, 28^ with complete genetic and clinical data were included in the survival analyses testing the association between RAG-mediated SV traits and patient outcomes. All data, including SV calls and breakpoint locations, were available in Pölönen et al. (2024). Analysis of RAG recombination at SV breakpoints was conducted for T-ALL using the same approach as described above for B-ALL patients. Consistent with prior studies ^28, 29^, multivariable Cox PH regression models with Firth's penalized likelihood method were fitted, adjusting for age at diagnosis, sex, WBC at diagnosis, genetic ancestry, molecular subtype, and day-29 MRD status. Molecular subtype was included as a binary covariate in survival analyses. Patients were grouped into high-risk or low-risk based on genetic subtypes in pediatric T-ALL patients as previously defined by Pölönen et al. ^28–30^.

**Supplemental Results**

### **RAG-mediated SVs and clinical outcomes in B-ALL patients**

Using the relapse as a binary outcome, we observed an increasing trend in relapse odds with increasing frequency of RAG-mediated SVs (p for non-linearity =0.052) (**Figure S11**). Consistent with time-to-event event-free survival for relapse results, we also observed the threshold effect that patients with $\geq$15 SVs (n=32) had a >5-fold increased risk of relapse (reference <median, HR: 5.21, 1.82-14.97). Excluding deletions, a relatively large group of patients (n=229) harboring 3 or more off-target RAG-mediated SVs had a >2-fold risk of relapse compared with patients with 0 non-deletion SVs (n=573) (OR: 2.13 [1.48, 3.06]). *ETV6::RUNX1* patients with 3 non-deletion SVs (n=126) had an ~3-fold risk of relapse compared with patients with 0 non-deletion SVs (n=154) (OR: 2.96 [1.68, 5.35]), and a similar pattern was seen in MRD-negative *ETV6::RUNX1* patients (OR: 3.77 [1.86, 7.95]).

For time-to-event outcomes including event-free survival (for relapse vs none), disease-free survival and overall survival, we observed similar increased risk for the total number of RAG-mediated SVs, off-target RAG-mediated SVs (RSS at at least one breakpoint or with only one RSS), particularly for non-deletion SVs (**Table S14**). These findings were further confirmed in the competing risk analysis for relapse risk.

### **RAG-mediated SVs and clinical outcomes in T-ALL patients**

To replicate findings from the B-ALL discovery cohort in T-ALL patients, we examined RAG-mediated SVs and the association with patient outcomes in an independent cohort of 1,328 T-ALL patients in COG trial AALL0434 ^28^. In contrast to MP2PRT, this T-ALL patient cohort was not selected to be enriched for relapsed cases. Of the 1,244 total patients, only n=123 (9.4%) experienced relapse events. A total of 6,972 SVs were identified among T-ALL patients (per patient median = 4 SVs, range: 0-69), of which 1,323 (19%) SVs had an RSS at ≥1 breakpoint and 1,072 (15.4%) SVs had RSS at only one breakpoint, consistent with aberrant RAG activity. Among off-target (non-Ig/TCR) SVs, 773 (12.3%) harbored RSS at only one breakpoint. Off-target RAG-mediated SVs with a single RSS were non-significantly associated with a higher risk of relapse (HR per 1 SV increase: 1.13, [0.94-1.33], p=0.178). Off-target RAG-mediated inversions with a single RSS were significantly associated with an ~50% increased relapse risk (HR: 1.46 [1.02-2.00], p=0.0383). This association was confirmed in Fine-Gray competing risk analyses accounting for non-relapse death (HR for relapse: 1.40, [1.02-1.93], p=0.036) (**Table S16**).
