## Supplementary Figures for "RAG-mediated structural variation and its impact on relapse risk in acute lymphoblastic leukemia"

#### Figure S1

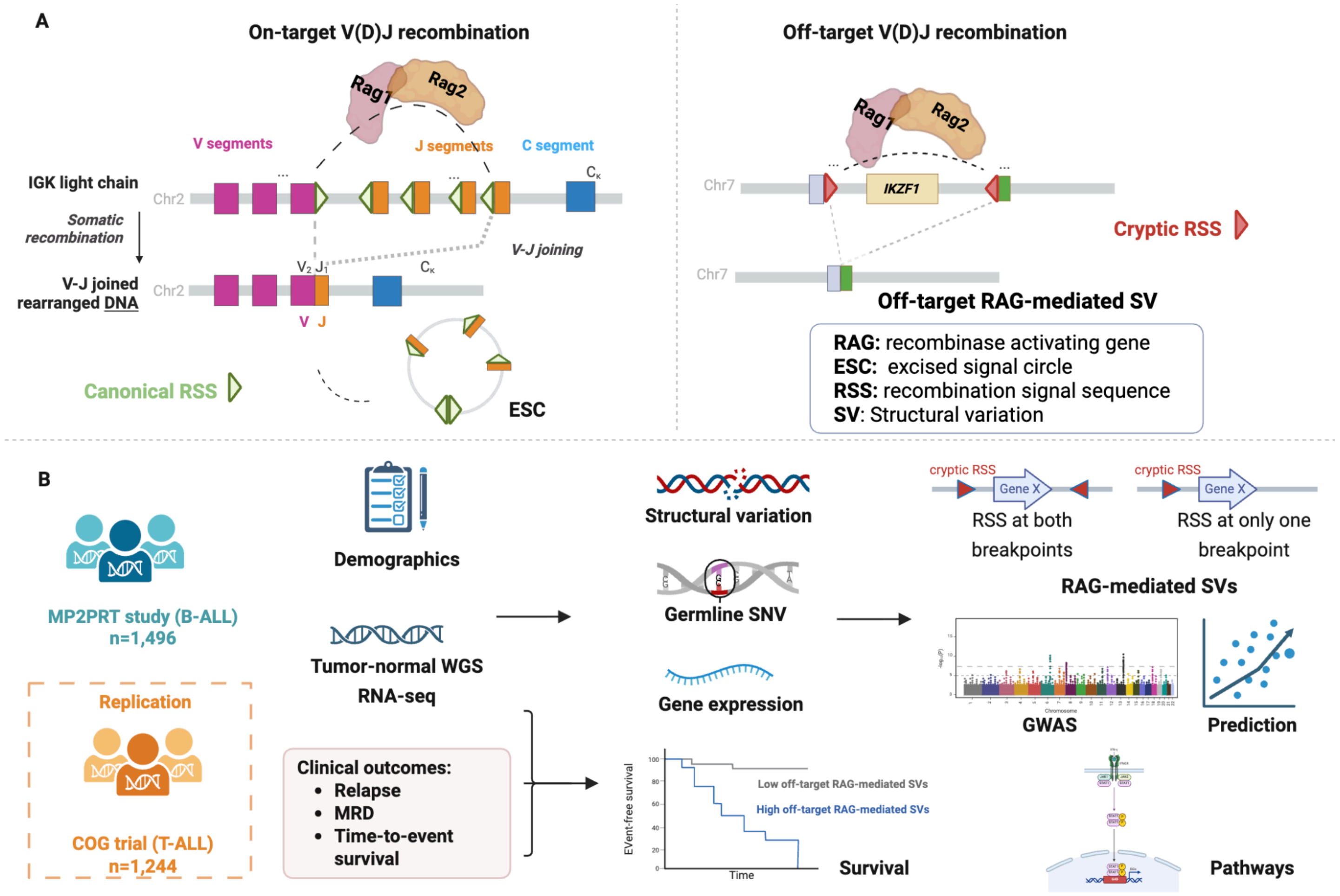


**Figure S1. V(D)J recombination and workflow of analysis.**

**A.** Graphical representation of on-target (left) and off-target (right) V(D)J or RAG recombination. On-target V(D)J recombination typically generates antibody diversity at the Ig and TCR gene regions, while off-target V(D)J recombination can result in structural variant (SV) formation that can drive leukemogenesis in ALL.

**B.** Workflow of analysis to investigate the association between RAG-mediated SVs, patient-level characteristics, and patient outcomes in the MP2PRT B-ALL cohort. Replication of the association between off-target RAG-mediated SVs and relapse risk was conducted in a COG T-ALL patient cohort. Figure was created using BioRender.com.

**Figure S2**

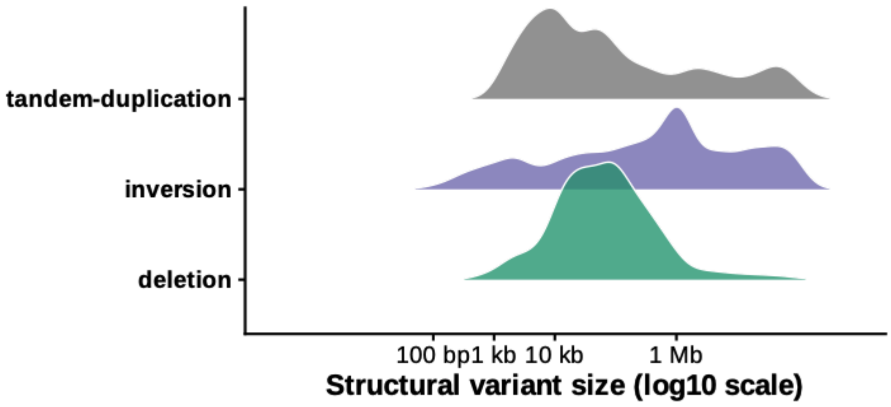


**Figure S2. Log-transformed size distributions of three major structural variant (SV) subtypes.** Density plots show the distribution of SV sizes (in base pairs) for deletions, inversions, and tandem duplications. The x-axis represents SV size on a log10 scale (ticks shown in bp, kb, and Mb), and the y-axis reflects relative density.

#### Figure S3

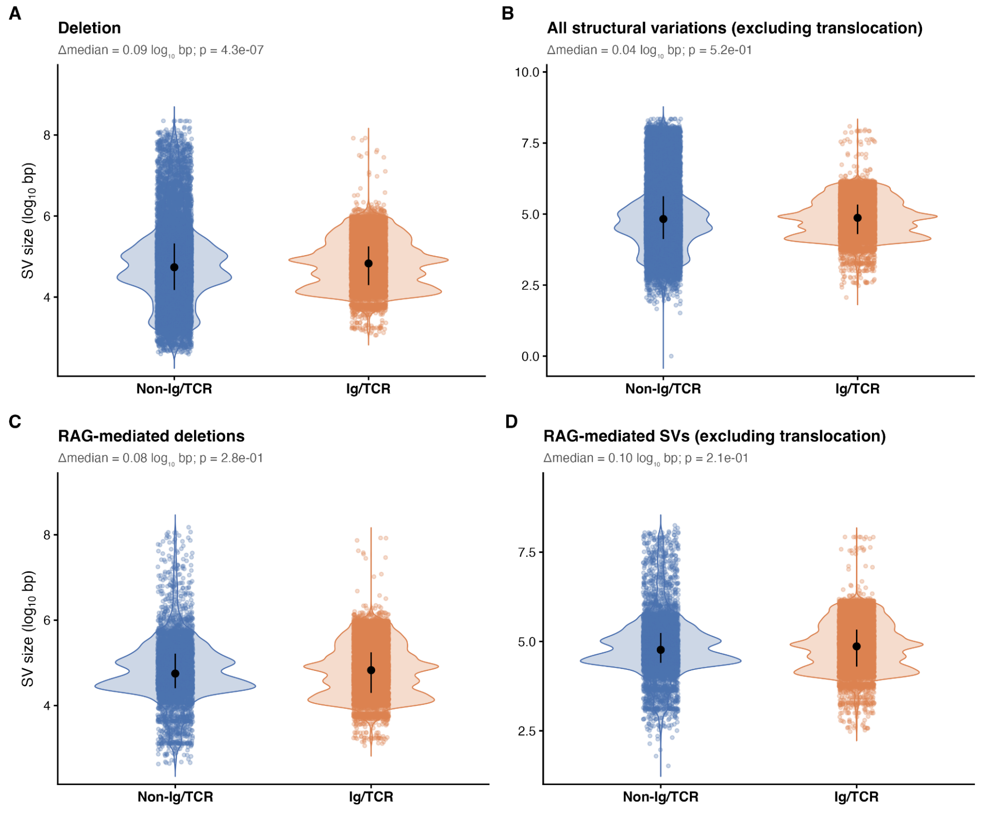


**Figure S3. Comparison of structural variant (SV) sizes between Non-Ig/TCR and Ig/TCR regions.** (**A**) Deletions. (**B**) All structural variants excluding translocations. (**C**).RAG-mediated deletions (**D**) RAG-mediated SVs excluding translocations. Violin plots display the distribution of SV sizes (log10-transformed base pairs) in Non-Ig/TCR and Ig/TCR regions. Points represent individual SV events; black dots indicate median values with vertical bars denoting interquartile ranges. The difference in median SV size (Δmedian, in log10 bp) and corresponding p-value (two-sided test) are shown above each panel.

#### Figure S4

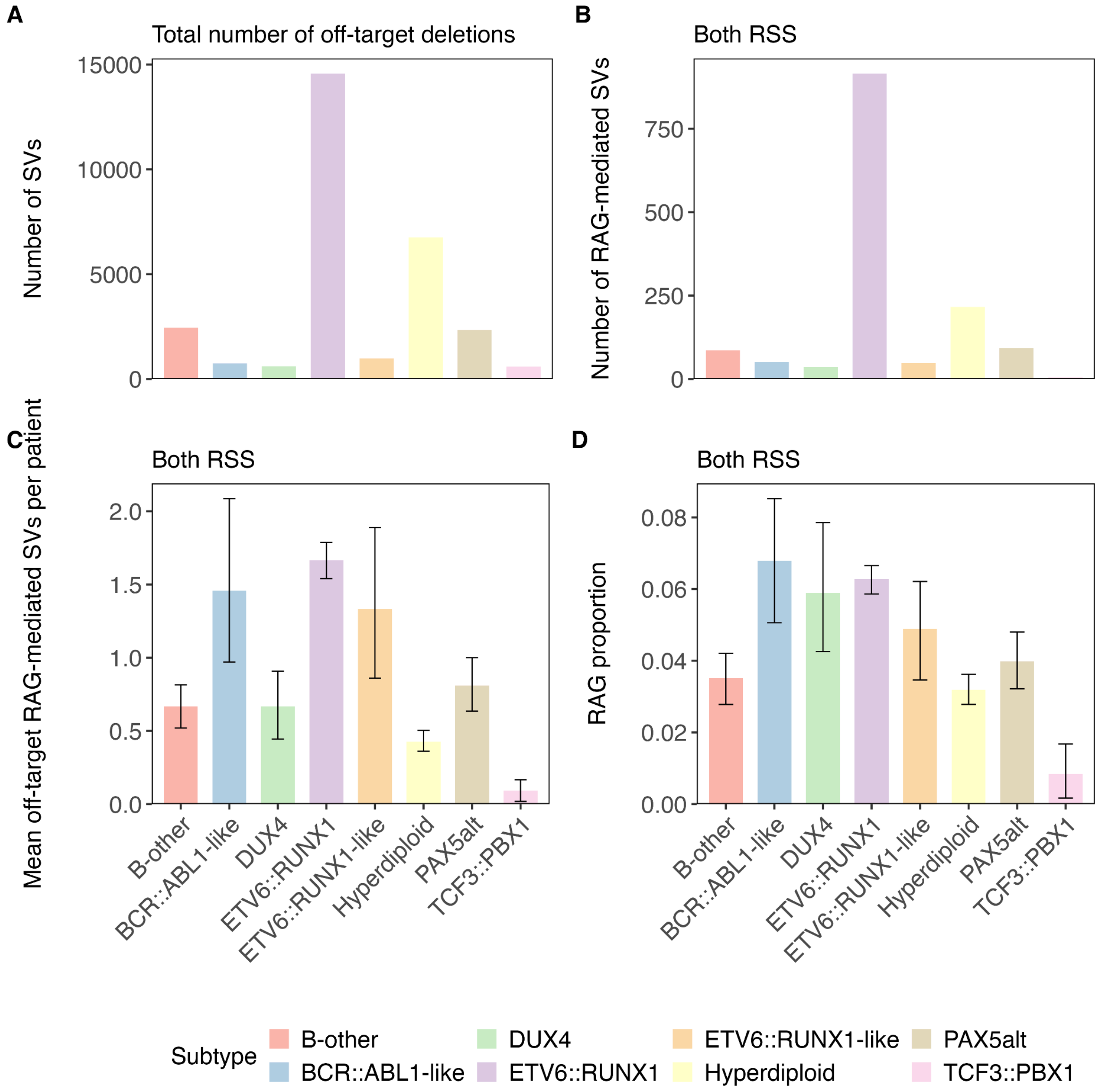


**Figure S4. Number and proportion of off-target RAG-mediated structural variations (SVs) (at both breakpoints) by B-cell ALL molecular subtype.**

Bar plots showing: (**A**) the number of total off-target (non-Ig/TCR) SVs, (**B**) the number of total off-target (non-Ig/TCR) RAG-mediated SVs (RSS at ≥1 breakpoint), (**C**) the average number of off-target RAG-mediated SVs per patient, and (**D**) the proportion of total off-target SVs that were RAG-mediated, across molecular subtypes of B-cell ALL in MP2PRT patients. Analyses were restricted to 1,479 patients with at least one off-target SVs, including B-other (n=129), BCR:ABL1-like (n=35), DUX4-rearranged (n=54), ETV6::RUNX1 (n=550), ETV6::RUNX1-like (n=36), Hyperdiploid (n=506), PAX5alt (n=115) and TCF3::PBX1 (n=54) subtypes. Error bars represent 95% bootstrap confidence intervals. Figure (**E**) and (**F**) show RAG1 and RAG2 expression across all 1465 participants with tumor transcriptome profiling data, including B-other (n=117), BCR:ABL1-like (n=36), DUX4-rearranged (n=54), ETV6::RUNX1 (n=552), ETV6::RUNX1-like (n=36), Hyperdiploid (n=501), PAX5alt (n=116) and TCF3::PBX1 (n=53) subtypes. Gene expression levels calculated as fragments per kilobase of transcript per million mapped reads (FPKM) using RNA-seq data where forward and reverse strands are not distinguished.

#### Figure S5

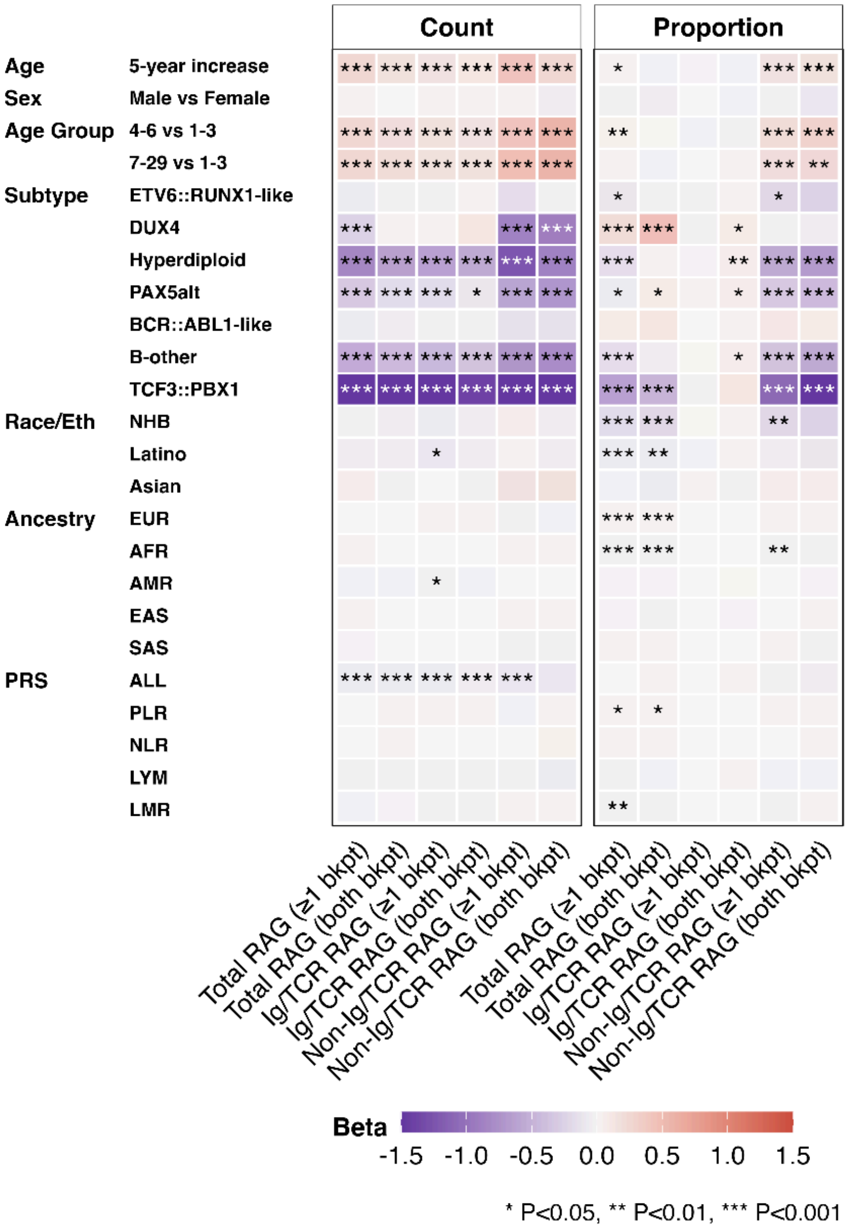


**Figure S5. Univariate associations of patient-level characteristics with proportion of RAG-mediated structural variation traits.** Univariate association results for structural variation (SV) proportion traits. The heatmap displays results from negative binomial models for SV count burden and from negative binomial models with offset (total SVs) for SV proportion burden. For subtype analysis, ETV6::RUNX1 subtype was the reference. For race/ethnicity (Race/Eth), non-Hispanic Whites were the reference, NHB = Non-Hispanic Black, Latino = Hispanic/Latino. Polygenic risk scores (PRS) were analyzed for ALL (acute lymphoblastic leukemia), platelet-to-lymphocyte ratio (PLR), neutrophil-to-lymphocyte ratio (NLR), lymphocyte counts (LYM), and lymphocyte-to-monocyte ratio (LMR). * P<0.05, ** P<0.01, *** P<0.001.

#### Figure S6

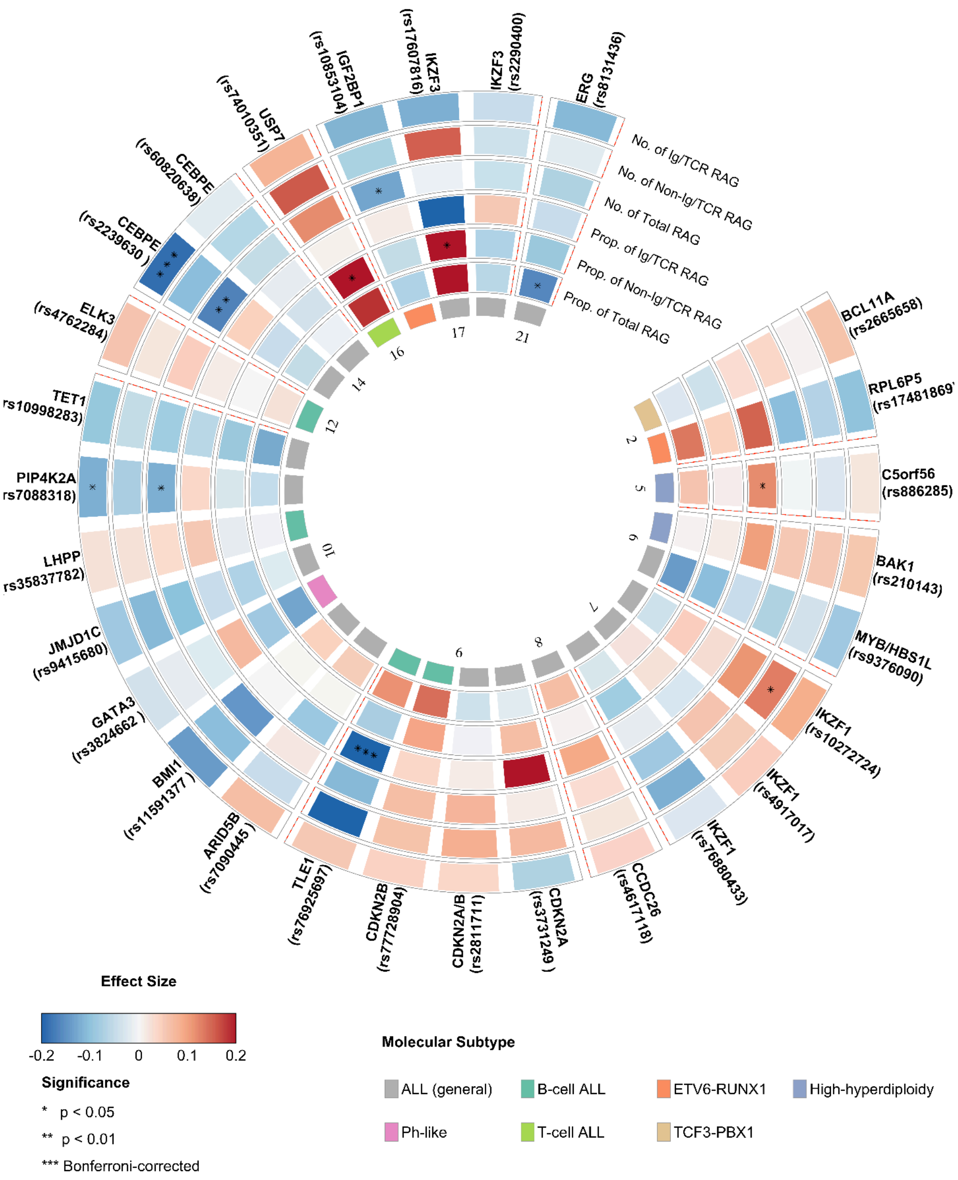


**Figure S6. Childhood ALL GWAS risk loci loci and RAG-mediated structural variation traits among 554 ETV6::RUNX1 patients.** Associations between 28 reported risk loci from GWAS of childhood acute lymphoblastic leukemia (ALL) and six RAG-mediated deletion traits (number/proportion of total/on-target/off-target RAG-mediated deletions(restricted to 554 ETV6::RUNX1 patients). We adjusted for age, sex, and the top 3 PCs . Genes and single nucleotide polymorphism (SNP) rsIDs are depicted in the radials with one band per phenotype, and divided by chromosomes. Colors indicate the magnitude and direction of the effect size. Gene–phenotype pairs reaching Bonferroni-corrected significance ($p < 0.001786$ (0.05/28)) are denoted by three asterisks.

#### Figure S7

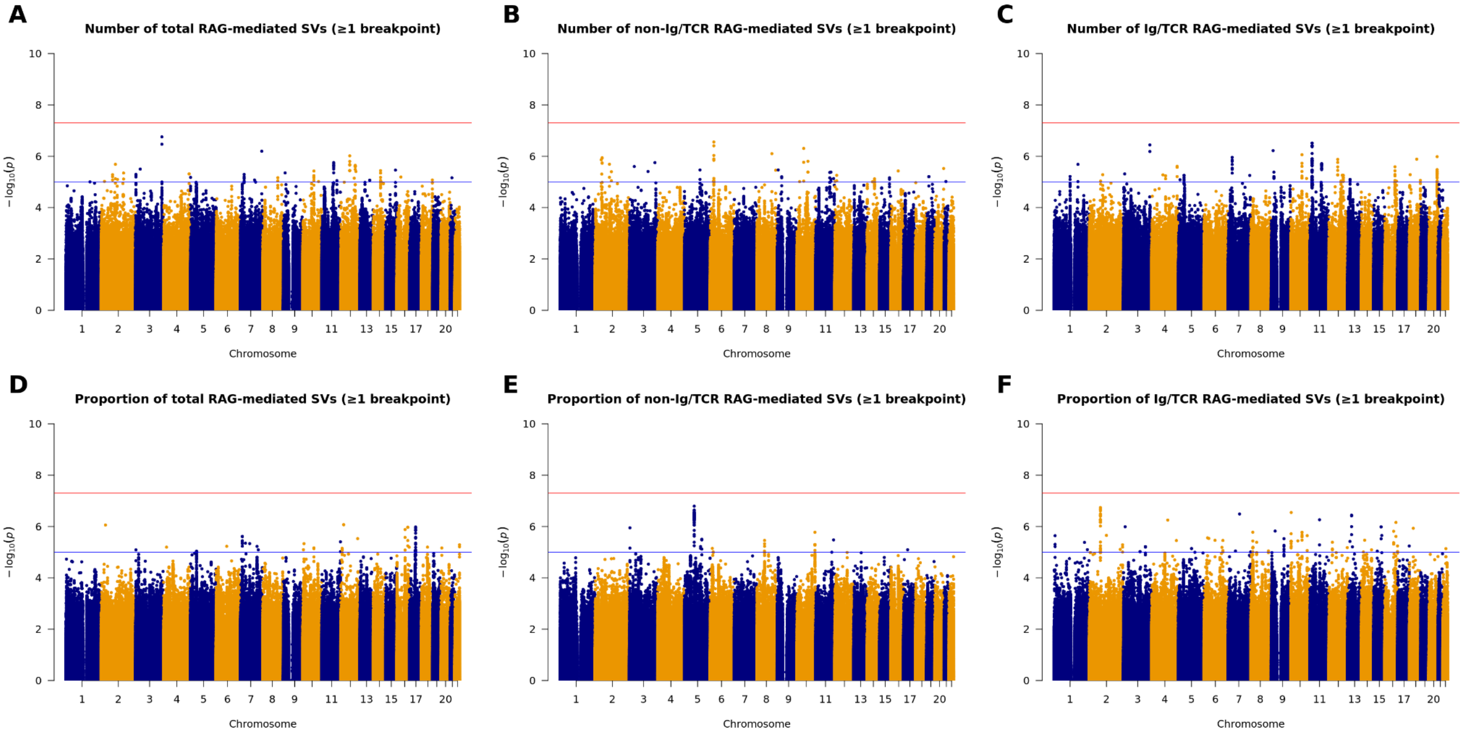


**Figure S7. GWAS results for RAG-mediated SV traits in B-ALL patients.**
Manhattan plots of GWAS result in the full multi-ancestry cohort. GWAS of number and proportion of total (overall), off-target and on-target RAG-mediated SVs (at least one breakpoint). Each panel displays genome-wide association results for a distinct SV outcome, with genomic position along the x-axis (ordered by chromosome) and −log10(p-values) on the y-axis. Points represent individual variants, colored alternately by chromosome for clarity. Two horizontal lines indicate genome-wide significance threshold ($P = 5 \times 10^{-8}$) and suggestive significance threshold ($P = 1 \times 10^{-5}$).

#### Figure S8

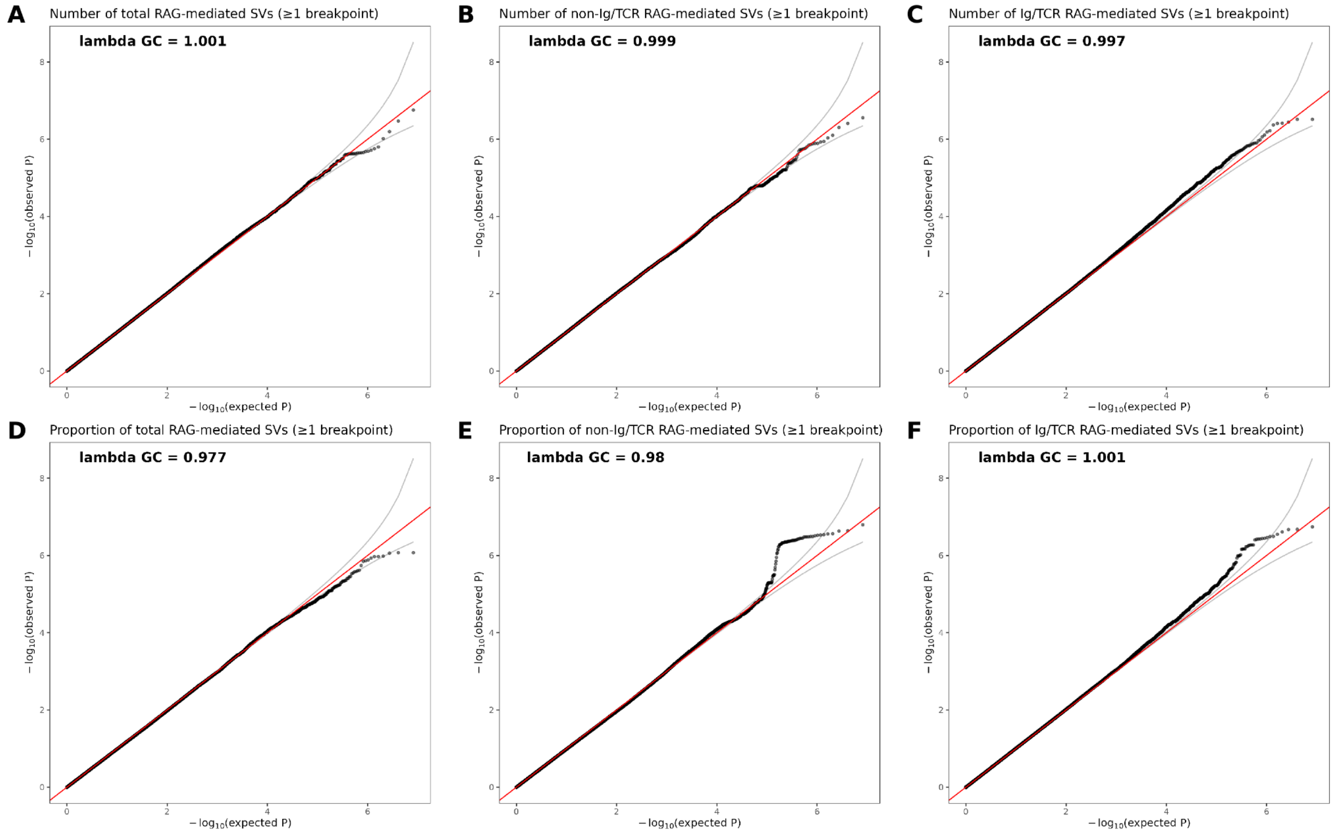


**Figure S8. Quantile–quantile (Q–Q) plots of RAG-mediated SV traits.** Quantile–quantile (Q–Q) plots comparing observed versus expected −log10(p-values) for deletion burden across different RSS event categories. Each panel corresponds to a distinct deletion outcome, as labeled. The red diagonal line indicates the null expectation under no association, and points represent observed test statistics. Genomic inflation factors (λ) are shown in each panel, indicating minimal deviation from the null and suggesting adequate control of test statistic inflation.

### Figure S9

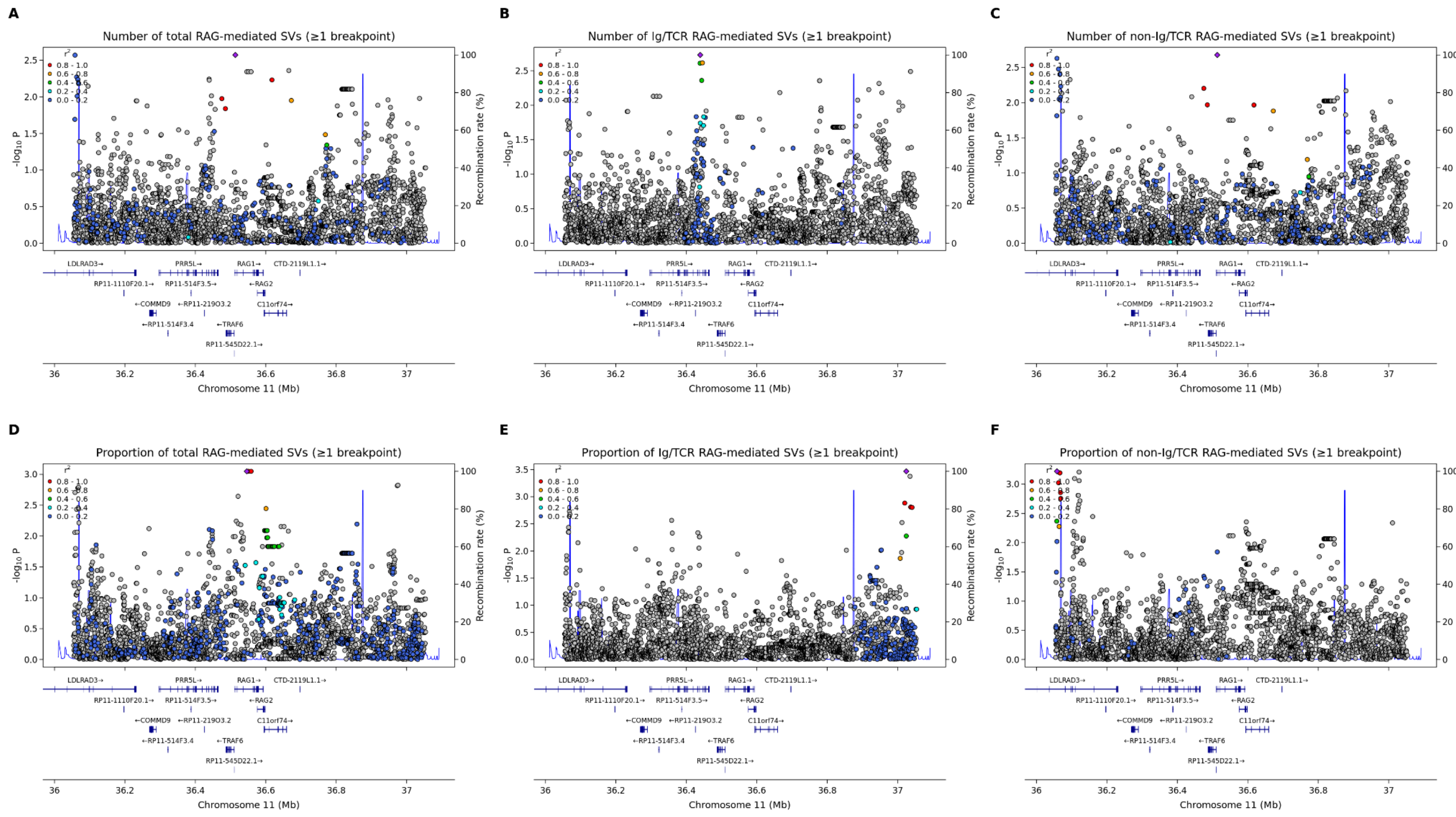


**Figure S9. Regional association plots for RAG-mediated SV traits at the chromosome 11 *RAG1*/*RAG2*.** Regional association plots showing association signals in chr11:36054295-37054295 for six SV-related phenotypes: (A) number of total RAG-mediated SVs (≥1 breakpoint), (B) number of Ig/TCR RAG-mediated SVs (≥1 breakpoint), (C) number of non-Ig/TCR RAG-mediated SVs (≥1 breakpoint), (D) proportion of total RAG-mediated SVs (≥1 breakpoint), (E) proportion of Ig/TCR RAG-mediated SVs (≥1 breakpoint), and (F) proportion of non-Ig/TCR RAG-mediated SVs (≥1 breakpoint). The lead variant in each panel is highlighted in red. Variants are additionally colored according to linkage disequilibrium with the lead variant, and the blue line indicates local recombination rate (right y-axis).

#### Figure S10

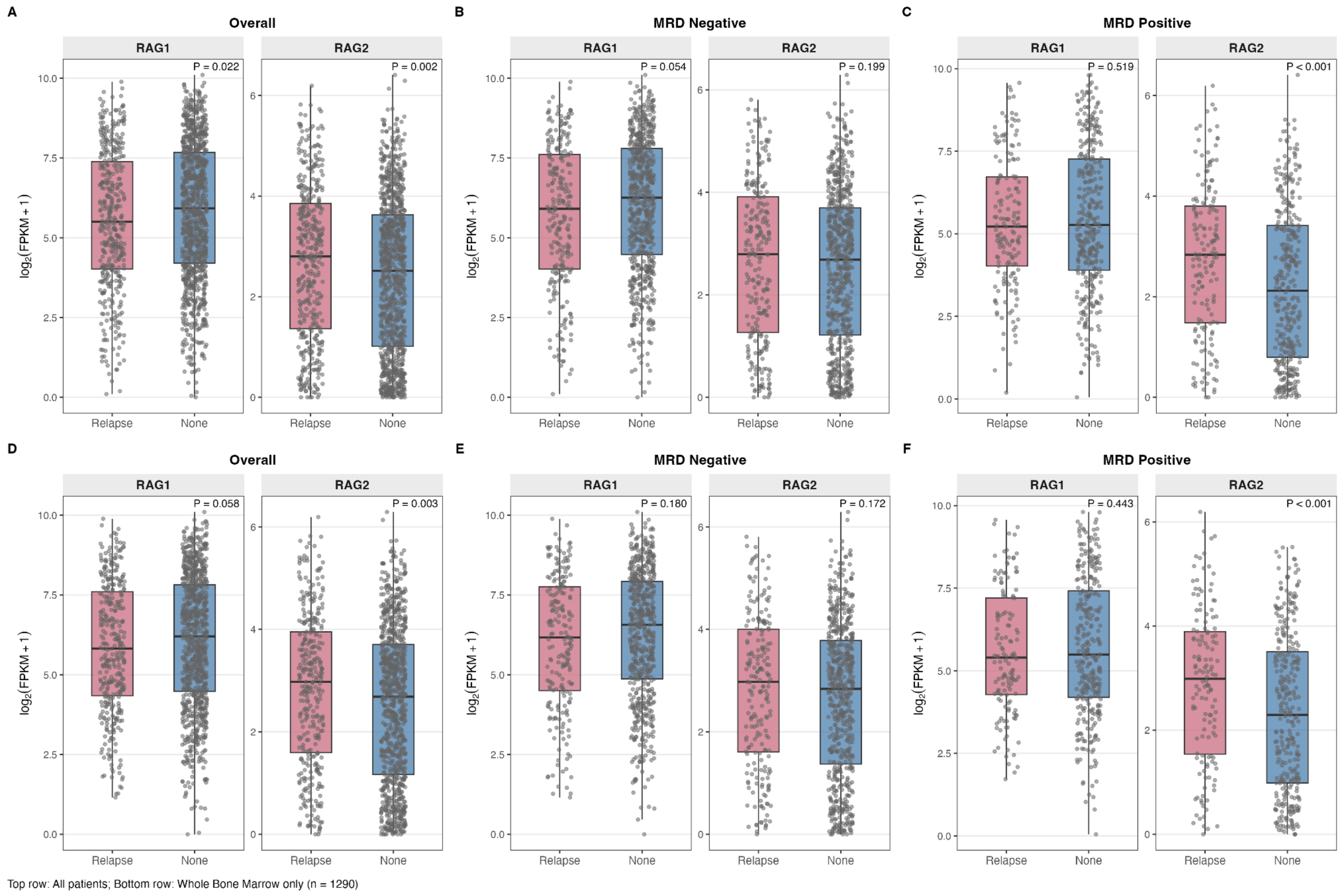


**Figure S10. RAG1/RAG2 expression by relapse status in MP2PRT B-ALL patients.**

Box and whisker plots displaying expression levels of RAG1 and RAG2 genes by relapse status in MP2PRT B-ALL patients overall, and by MRD status. Panels A-C display results across all 1,465 patients with tumor transcriptome profiling data, including n=177 with peripheral whole blood specimens and n=1,288 with whole bone marrow specimens. Panels D-F display results restricted to the n=1,288 participants with whole bone marrow specimens. Gene expression levels were calculated as fragments per kilobase of transcript per million mapped reads (FPKM) using RNA-seq data where forward and reverse strands were not distinguished.

**Figure S11**

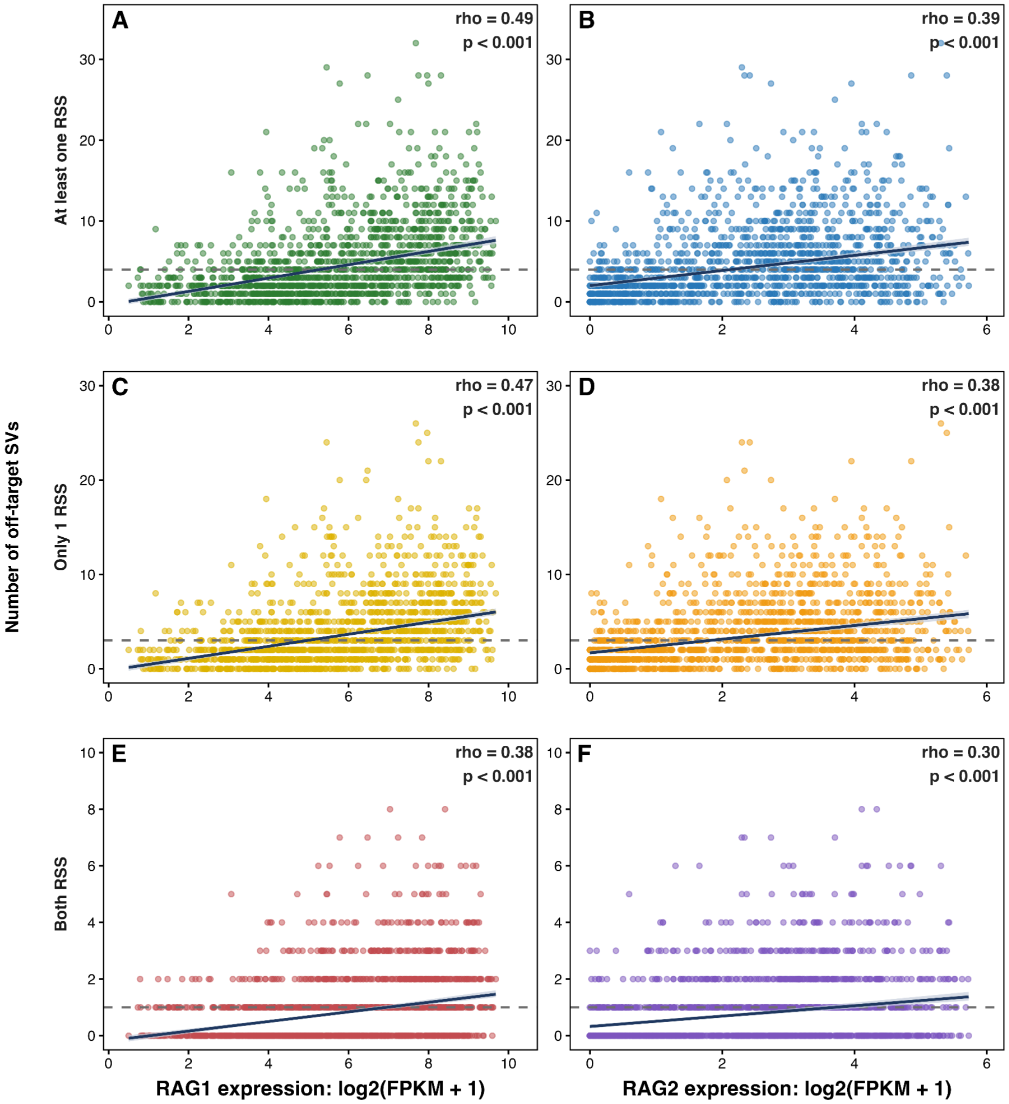


**Figure S11. Association between RAG1/RAG2 expression and off-target structural variation burden.** Scatter plots show the relationship between RAG1/RAG2 expression level (log2(FPKM+1) and the number of off-target SVs (at least one RSS, only one RSS, both RSS) across samples. Solid lines indicated fitted trends with shaded confidence intervals. Spearman's rank correlation coefficient (rho) and *p* value are reported for each comparison, demonstrating a positive association between RAG1 /RAG2 expression and off-target SV counts across all RSS enrichment categories.

**Figure S12**

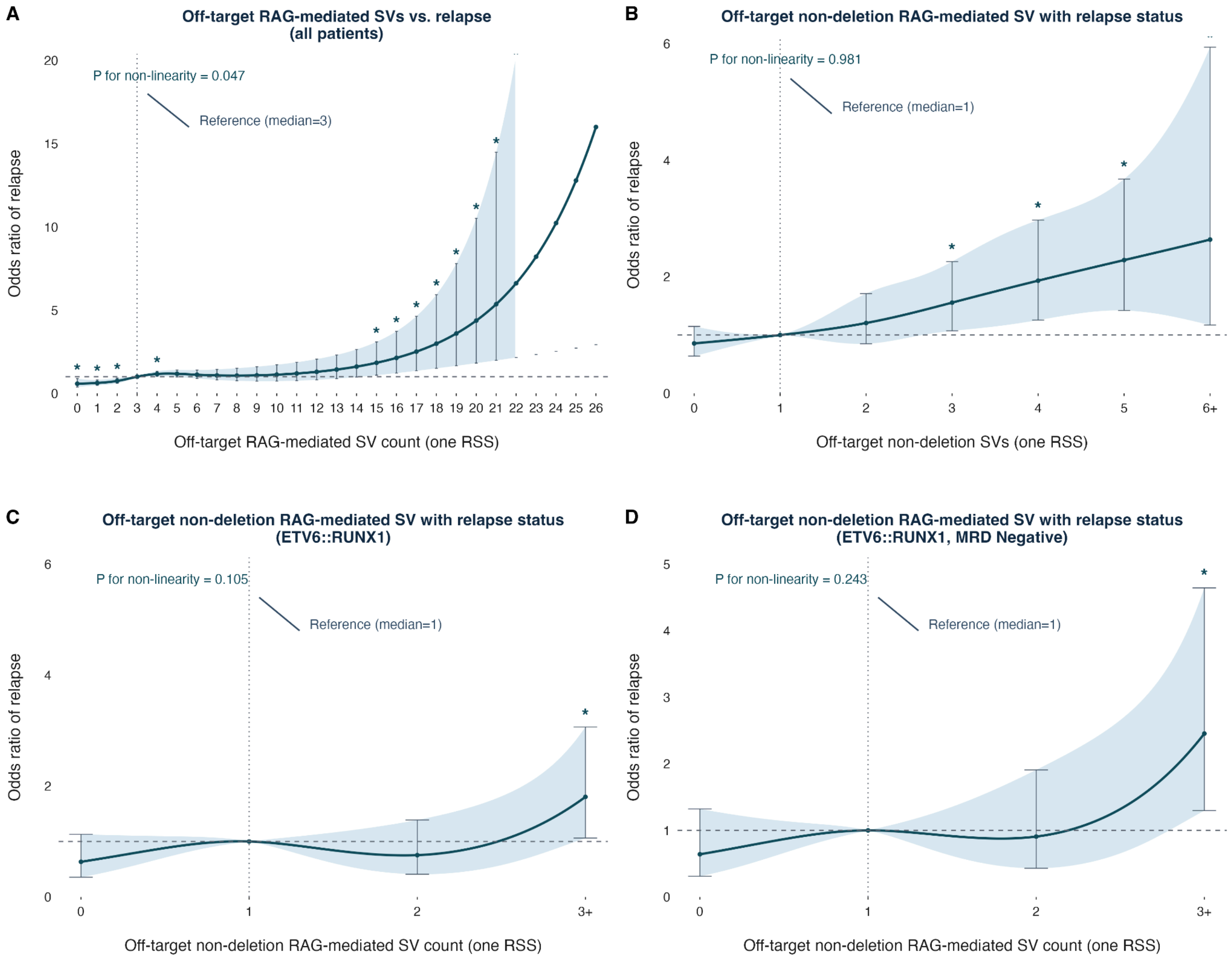


**Figure S12. Off-target structural variation burden and relapse risk across different SV count strata.** Association between off-target RAG-mediated SV count with one RSS and odds ratio (OR) of relapse.Relapse risk was modeled as a binary outcome in the multivariable logistic regression model with an additional natural cubic spline term for the off-target SV count (one RSS), adjusting for age (per 5-year increase), sex, race/ethnicity and molecular subtype. Likelihood ratio test was used to test for non-linearity. The solid curve represents estimated OR relative to the cohort median SV count (reference), and the shaded band indicates the 95% confidence interval. The vertical dotted line and annotation mark the median reference value. **A**. Relapse risk and off-target RAG-mediated SVs (only one RSS). **B**. Relapse risk and off-target RAG-mediated non-deletion SVs (only one RSS). **C**. Relapse risk and off-target RAG-mediated non-deletion SVs (only one RSS) among ETV6::RUNX1 patients. **D**. Relapse risk and off-target RAG-mediated non-deletion SVs (only one RSS) among ETV6::RUNX1 MRD-negative patients.

### Figure S13

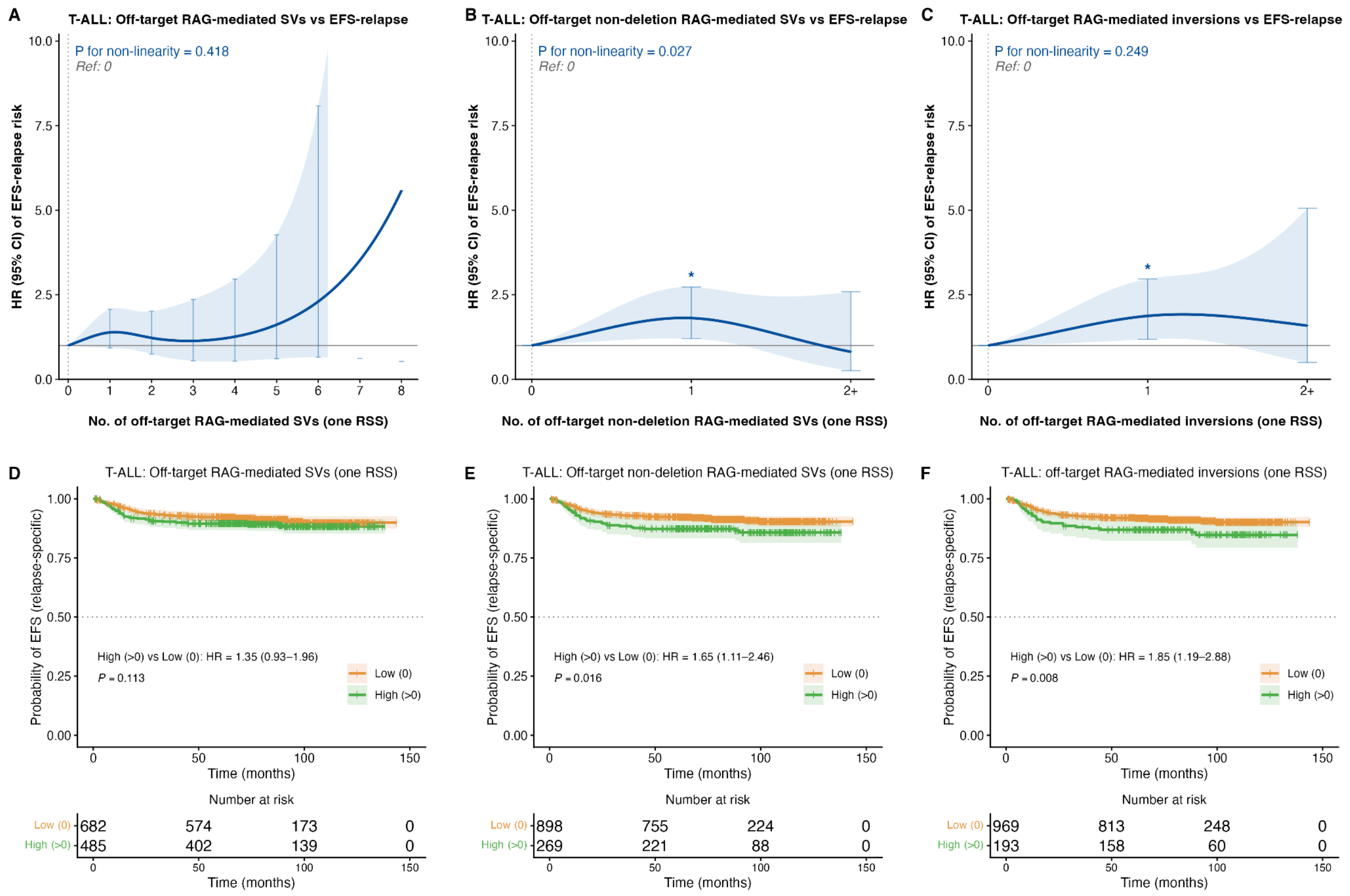


**Figure S13. Association between off-target RAG-mediated SVs and relapse risk in T-cell ALL patients.** **A**. Association between off-target RAG-mediated SVs with only one RSS and hazard ratio (HR) of event-free survival for relapse in T-ALL patients. Multivariable penalized Cox proportional hazards model was fitted, adjusting for age at diagnosis, sex, WBC at diagnosis, genetic ancestry, molecular subtype, and day-29 MRD status. P for non-linearity was assessed by the likelihood ratio test. A. Relapse risk and off-target RAG-mediated SVs (only one RSS). **B**. Relapse risk and off-target RAG-mediated non-deletion SVs (only one RSS). C. Relapse risk and off-target RAG-mediated inversions (only one RSS). **D**. Kaplan-Meier curves for event-free survival (EFS) for relapse in patients stratified by off-target RAG-mediated SV burden (High>0) versus low (=0) (with one RSS). **E**. Kaplan-Meier curves for EFS in patients stratified by off-target RAG-mediated non-deletion SVs (with one RSS). **F**. Kaplan-Meier curves for EFS in patients stratified by off-target RAG-mediated inversions (with one RSS).
